# A global atlas of genetic associations of 220 deep phenotypes

**DOI:** 10.1101/2020.10.23.20213652

**Authors:** Saori Sakaue, Masahiro Kanai, Yosuke Tanigawa, Juha Karjalainen, Mitja Kurki, Seizo Koshiba, Akira Narita, Takahiro Konuma, Kenichi Yamamoto, Masato Akiyama, Kazuyoshi Ishigaki, Akari Suzuki, Ken Suzuki, Wataru Obara, Ken Yamaji, Kazuhisa Takahashi, Satoshi Asai, Yasuo Takahashi, Takao Suzuki, Nobuaki Shinozaki, Hiroki Yamaguchi, Shiro Minami, Shigeo Murayama, Kozo Yoshimori, Satoshi Nagayama, Daisuke Obata, Masahiko Higashiyama, Akihide Masumoto, Yukihiro Koretsune, Kaoru Ito FinnGen, Chikashi Terao, Toshimasa Yamauchi, Issei Komuro, Takashi Kadowaki, Gen Tamiya, Masayuki Yamamoto, Yusuke Nakamura, Michiaki Kubo, Yoshinori Murakami, Kazuhiko Yamamoto, Yoichiro Kamatani, Aarno Palotie, Manuel A. Rivas, Mark J. Daly, Koichi Matsuda, Yukinori Okada

## Abstract

Current genome-wide association studies (GWASs) do not yet capture sufficient diversity in populations and scope of phenotypes. To expand an atlas of genetic associations in non-European populations, we conducted 220 deep-phenotype GWASs (diseases, biomarkers, and medication usage) in BioBank Japan (*n*=179,000), by incorporating past medical history and text-mining of electronic medical records. Meta-analyses with the UK Biobank and FinnGen (*n*_total_=628,000) identified ∼5,000 novel loci, which improved the resolution of genomic map of human traits. This atlas elucidated landscape of pleiotropy as represented by MHC locus, where we conducted HLA fine-mapping. Finally, we performed statistical decomposition of matrices of phenome-wide summary statistics, and identified latent genetic components, which pinpointed responsible variants and biological mechanisms underlying current disease classifications across populations. The decomposed components enabled genetically-informed subtyping of similar diseases (e.g., allergic diseases). Our study suggests a potential avenue for hypothesis-free re-investigation of human diseases through genetics.

## Introduction

Medical diagnosis has been shaped through description of organ dysfunctions and extraction of shared key symptoms, which categorizes a group of individuals into a specific disease to provide an optimal treatment. The earliest physicians in ancient Egypt empirically made disease diagnoses based on clinical symptoms, palpitation, and auscultation (∼2600 BC)^1^. Since then, physicians have refined the disease diagnosis by empirically categorizing the observed symptoms (e.g., cough, sputum, and fever) to describe underlying dysfunction (e.g., pneumonia, a lung infection). An increased understanding of organ functions and the availability of diagnostic tests including biomarkers and imaging techniques have contributed to the current disease classifications, such as ICD10^2^ and phecode^3^.

In the past decades, genome-wide association studies (GWASs)^4^ and phenome-wide association studies (PheWASs)^5^ have provided novel insights into the biological basis underlying disease diagnoses. While disease pathogenesis is quite multifactorial, genetic underpinnings provide us with one way to independently assess the validity of historically-defined disease classifications. To this end, a comprehensive catalog of disease genetics is warranted. While previous works have broadly contributed the catalog^6^, current genetic studies are still short of comprehensiveness in three ways; (i) population, in that the vast majority of GWASs have been predominated by European populations^7^, (ii) scope of phenotypes, which were mostly limited to pre-determined diseases on which participants’ recruitment had been performed, and (iii) a systematic method to interpret a plethora of summary results for understanding disease pathogenesis and epidemiology. We thus need to promote equity in genetic studies by sharing the results of genetic studies of deep phenotypes from diverse populations.

To expand the atlas of genetic associations, here we conducted 220 deep-phenotype GWASs in BioBank Japan (BBJ), including 108 phenotypes on which GWAS has never been conducted in East Asian populations. We then conducted GWASs for corresponding phenotypes in UK Biobank (UKB) and FinnGen, and performed cross-population meta-analyses (*n*_total_=628,000). We sought to elucidate the landscape of pleiotropy and genetic correlation across diseases and populations. Furthermore, we applied DeGAs^8^ to perform truncated singular-value decomposition (TSVD) on matrices of GWAS summary statistics of 159 diseases each in Japanese and European ancestries, and derived latent components shared across the diseases. We interpreted the derived components by (i) functional annotation of genetic variants explaining the component, (ii) identification of important cell types where the genes contributing to each component are specifically regulated, and (iii) projection of GWASs of biomarkers or metabolomes into the component space. The latent components recapitulated the hierarchy of current disease classifications, while different diseases sometimes converged on the same component which implicated shared biological pathways and relevant tissues. We classified a group of similar diseases (e.g., allergic diseases) into subgroups based on these components. Analogous to the conventional classification of diseases structured by the shared symptoms, an atlas of genetic studies suggested the latent structure behind human diseases, which can elucidate the genetic variants, genes, organs, and biological functions underlying human diseases.

## Results

### GWAS of 220 traits in BBJ and cross-population meta-analysis

Overview of this study is presented in **Extended Data Figure 1**. BBJ is a nationwide biobank in Japan, and recruited participants based on the diagnosis of at least one of 47 target diseases (**Supplementary Note**)^9^. Along with the target disease status, deep phenotype data, such as past medical history (PMH), drug prescription records (∼7 million), text data retrieved from electronic medical records (EMR), and biomarkers, have been collected. Beyond the collection of case samples based on the pre-determined target diseases, the PMH and EMR have provided broader insights into disease genetics, as shown in recently launched biobanks such as UKB^10^ and BioVU^11^. We therefore curated the PMH, performed text-mining of the EMR, and merged them with 47 target disease status.^12^ We created individual-level phenotypes on 159 disease endpoints (38 target diseases with median 1.25 times increase in case samples and 121 novel disease endpoints) and 23 categories of medication usage. We then systematically mapped the disease endpoints into phecode and ICD10, to enable harmonized GWASs in UKB and FinnGen. We also analyzed a quantitative phenotype of 38 biomarkers in BBJ, of which individual phenotype data are available in UKB^13^. Using genotypes imputed with the 1000 Genome Project phase 3 data (*n=*2,504) and population-specific whole-genome sequencing data (*n*=1,037) as a reference panel^14^, we conducted the GWASs of 159 binary disease endpoints, 38 biomarkers, and 23 medication usages in ∼179,000 individuals in BBJ (**Figure 1a–c**, **Supplementary Table 1** and **2** for phenotype summary). To maximize statistical power, we used a linear mixed model implemented in SAIGE^15^ for binary traits and BOLT^16^ for quantitative traits. By using linkage disequilibrium (LD)-score regression^17^, we confirmed that potential biases were controlled in the GWASs (**Supplementary Table 3**). In this expanded scope of GWASs in the Japanese population, we identified 519 genome-wide significant loci across 159 disease endpoints, 2,249 across 38 biomarkers, of which 113 and 281 loci were novel, respectively (*P<*5.0×10^-8^; see **Methods**, **Supplementary Table 4**). We conducted the initial medication-usage GWASs in East Asian populations, and detected 215 genome-wide significant loci across 23 traits (see **Methods**). These signals underscore the value of (i) conducting GWASs in non-Europeans and (ii) expanding scope of phenotypes by incorporating biobank resources such as PMH and EMR. For example, we detected an East Asian-specific variant, rs140780894, at the MHC locus in pulmonary tuberculosis (PTB; Odds Ratio [OR]=1.2, *P*=2.9×10^-23^, Minor Allele Frequency[MAF]_EAS_=0.24; **Extended Data Figure 2**), which was not present in European population (Minor Allele Count [MAC]_EUR_=0)^18^. PTB is a serious global health burden and relatively endemic in Japan^19^ (annual incidence per 100,000 was 14 in Japan whereas 8 in the United Kingdom and 3 in the United States in 2018 [World Health Organization, Global Tuberculosis Report]). Because PTB, an infectious disease, can be treatable and remittable, we substantially increased the number of cases by combining the participants with PMH of PTB to the patients with active PTB at the time of recruitment (from 549^12^ to 7,800 case individuals). We also identified novel signals in common diseases that had not been target diseases but were included in the PMH record, such as rs715 at 3’UTR of *CPS1* in cholelithiasis (**Extended Data Figure 3**; OR=0.87, *P*=9.6×10^-13^) and rs2976397 at the *PSCA* locus in gastric ulcer, gastric cancer, and gastric polyp (**Extended Data Figure 4**; OR=0.86, *P*=6.1×10^-24^ for gastric ulcer). We detected pleiotropic functional variants, such as a deleterious missense variant, rs28362459 (p.Leu20Arg), in *FUT3* associated with gallbladder polyp (OR=1.46, *P*=5.1×10^-11^) and cholelithiasis (OR=1.11, *P*=7.3×10^-9^; **Extended Data Figure 5**), and a splice donor variant, rs56043070 (c.89+1G>A), causing loss of function of *GCSAML* associated with urticaria (OR=1.24, *P*=6.9×10^-12^; **Extended Data Figure 6**), which was previously reported to be associated with platelet and reticulocyte counts^4^. Medication-usage GWASs also provided interesting signals as an alternative perspective for understanding disease genetics^20^. For example, individuals taking HMG CoA reductase inhibitors (C10AA in Anatomical Therapeutic Chemical Classification [ATC]) were likely to harbor variations at *HMGCR* (lead variant at rs4704210, OR=1.11, *P*=2.0×10^-27^). Prescription of salicylic acids and derivatives (N02BA in ATC) were significantly associated with a rare East Asian missense variant in *PCSK9*, rs151193009 (p.Arg93Cys; OR=0.75, *P*=7.1×10^-11^, MAF_EAS_=0.0089, MAF_EUR_=0.000; **Extended Data Figure 7**), which might indicate a strong protective effect against thromboembolic diseases in general.

**Figure 1.**
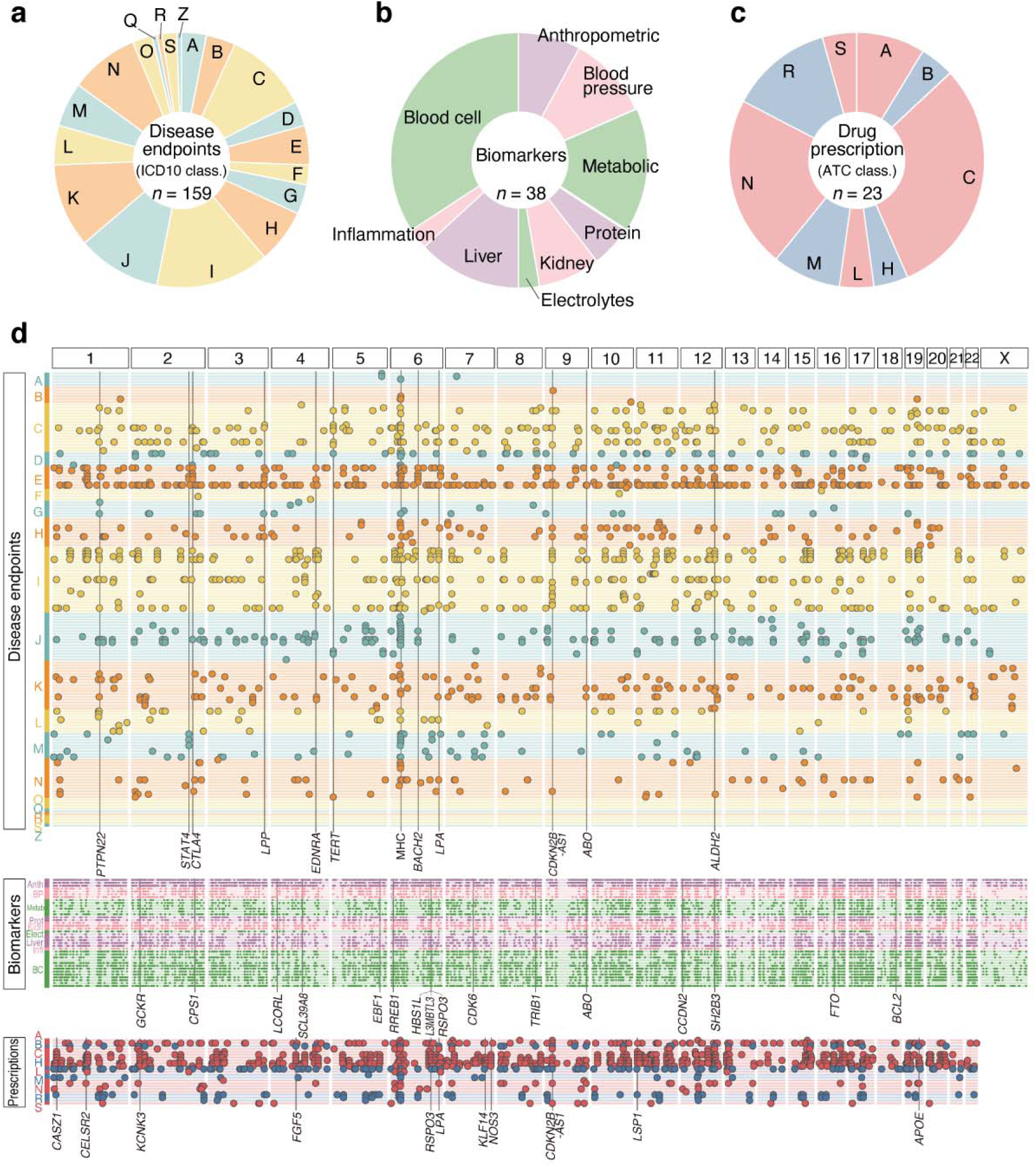
Overview of the identified loci in the cross-population meta-analyses of 220 deep phenotype GWASs. (**a**-**c**) The pie charts describe the phenotypes analyzed in this study. The disease endpoints (**a**; *n*_trait_=159) were categorized based on the ICD10 classifications (A to Z; **Supplementary Table 1a**), the biomarkers (**b**; *n*_trait_=38; **Supplementary Table 1b**) were classified into nine categories, and medication usage was categorized based on the ATC system (A to S; **Supplementary Table 1c**). (**d**) The genome-wide significant loci identified in the cross-population meta-analyses and pleiotropic loci (*P<*5.0×10^-8^). The traits (rows) are sorted as shown in the pie chart, and each dot represents significant loci in each trait. Pleiotropic loci are annotated by lines with a locus symbol.

We next conducted GWASs of corresponding phenotypes (i.e., disease endpoints and biomarkers) which can be mapped in UKB and FinnGen (196 and 128 traits, respectively; see **Methods**), and collected summary statistics of medication usage GWAS conducted in UKB^20^ (23 traits; **Supplementary Table 5**). To confirm that the signals identified in BBJ were reasonable, we systematically compared the effect sizes of the genome-wide significant variants in BBJ with those in a European dataset across binary and quantitative traits (see **Methods**). The loci identified in BBJ GWASs were successfully validated in the same effect direction (2,171 out of 2,305 [94.2%], *P*<10^-325^ in sign test) and with high effect-size correlation (**Extended Data Figure 8**). We also note that the genetic correlations encompassing genome-wide polygenic signals were generally high between BBJ and European GWASs (median ρ_ge_=0.82; **Supplementary Table 6**, see **Methods**).

Motivated by the high replicability, we performed cross-population meta-analyses of these 220 harmonized phenotypes across three biobanks (see **Methods**). We identified 1,730 disease-associated, 12,066 biomarker-associated, and 1,018 medication-associated loci in total, of which 571, 4,471, and 301 were novel, respectively (**Figure 1d**, **Supplementary Table 7**). We note that when we strictly control for multiple testing burden by Bonferroni correction (*P<*5.0×10^-8^ / (220 phenotypes × 3 populations)=7.6×10^-11^), the number of significantly associated loci was 844, 7,309, and 500, respectively. All these summary statistics of GWASs are openly distributed through PheWeb.jp website, with interactive Manhattan plots, Locus Zoom plots, and PheWAS plots based on PheWeb platform^21^. Together, we successfully expanded the genomic map of human complex traits in terms of populations and scope of phenotypes through conducting deep-phenotype GWASs across cross-population nationwide biobanks.

### The regional landscape of pleiotropy

Because human traits are highly polygenic and the observed variations within human genome are finite in number, pleiotropy, where a single variant affects multiple traits, is pervasive^22^. While pleiotropy has been intensively studied in European populations by compiling previous GWASs^22–25^, the landscape of pleiotropy in non-European populations has been understudied. By leveraging this opportunity for comparing the genetics of deep phenotypes across populations, we sought to investigate the landscape of regional pleiotropy in both Japanese and European populations. We defined the degree of pleiotropy as the number of significant associations per variant (*P*<5.0×10^-8^)^23^. In the Japanese, rs671, a missense variant at the *ALDH2* locus, harbored the largest number of genome-wide significant associations (47 traits; **Figure 2a**). Following this, rs2523559 at the MHC locus (24 traits) and rs1260326 at the *GCKR* locus (20 traits) were most pleiotropic. In Europeans, rs9265949 at the MHC locus harbored the largest number of genome-wide significant associations (46 traits; **Figure 2b**), followed by rs7310615 at the *ATXN2*/*SH2B3* locus (38 traits), rs1260326 at the *GCKR* locus (28 traits), and rs2519093 at *ABO* locus (28 traits). We note that those pleiotropic loci were not affected when we adjusted for phenotypically closely correlated traits (**Extended Data Figure 9,10**) or genetically closely related traits (**Extended Data Figure 10**, methods in **Supplementary Note**). Notably, the *ALDH2* locus (pleiotropic in Japanese) and the MHC locus (pleiotropic in Japanese and Europeans) are known to be under recent positive selection^26, 27^. To systematically assess whether pleiotropic regions in the genome were likely to be under selection pressure in each of the populations, we investigated the enrichment of the signatures of recent positive selection quantified by the metric singleton density score (SDS)^26^ values within the pleiotropic loci (see **Methods**).

**Figure 2.**
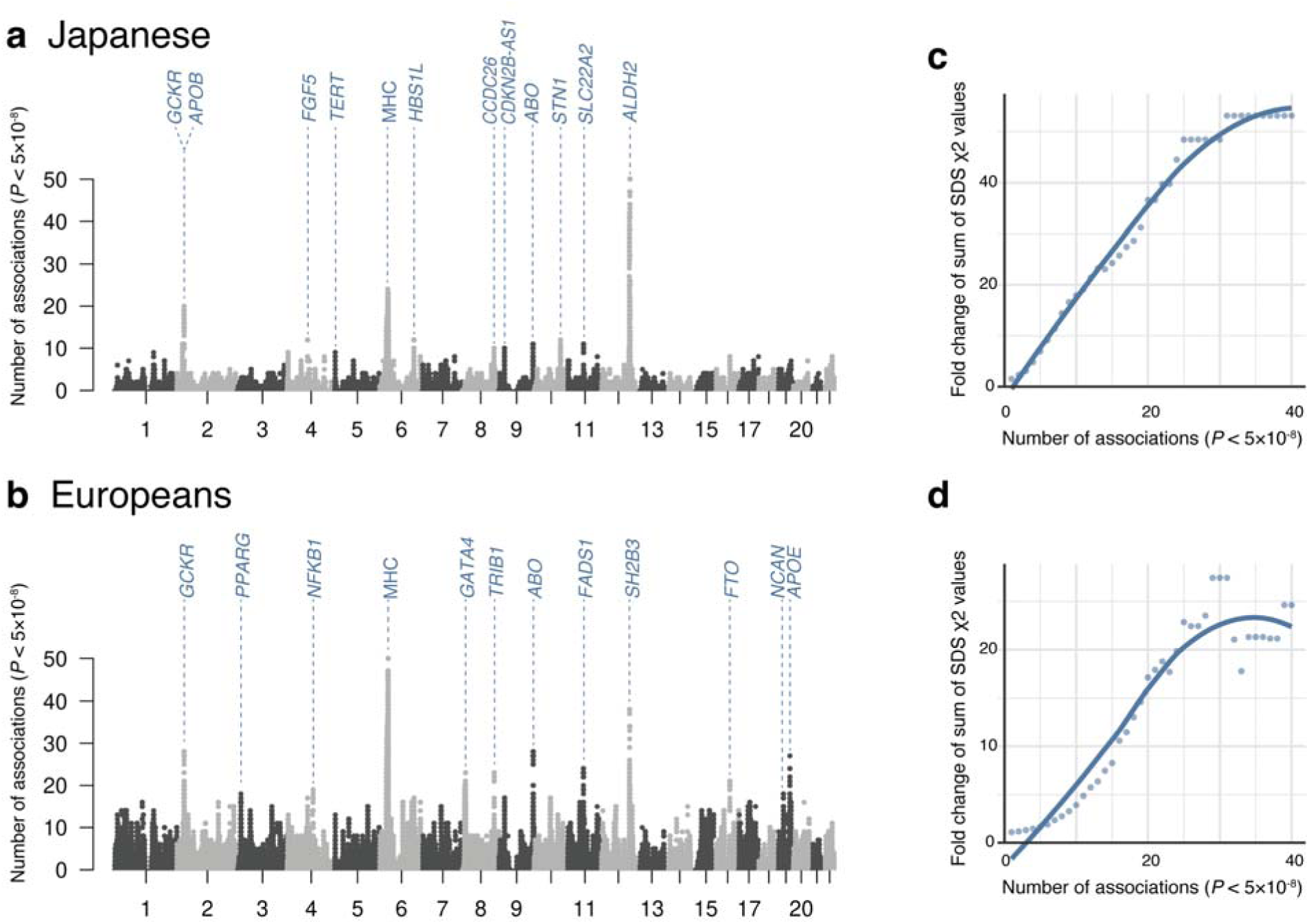
Number of significant associations per variant. (**a**, **b**) The Manhattan-like plots show the number of significant associations (*P*<5.0×10^-8^) at each tested genetic variant for all traits (*n*_trait_=220) in Japanese (**a**) and in European GWASs (**b**). Loci with a large number of associations were annotated based on the closest genes of each variant. (**c**, **d**) The plots indicate the fold change of the sum of SDS χ^2^ within variants with a larger number of significant associations than a given number on the x-axis compared with those under the null hypothesis in Japanese (**c**) and in Europeans (**d**). We also illustrated a regression line based on local polynomial regression fitting.

Intriguingly, when compared with those under the null hypothesis, we observed significantly higher values of SDS χ^2^ values within the pleiotropic loci, and this fold change increased as the number of associations increased (i.e., more pleiotropic) in both Japanese and Europeans (**Figure 2c** and **2d**). To summarize, the cross-population atlas of genetic associations elucidated the broadly shared landscape of pleiotropy, which implied a potential connection to natural selection signatures affecting human populations.

### Pleiotropic associations in HLA and ABO locus

Given the high degree of pleiotropy in both populations, we next sought to fine-map the pleiotropic signals within the MHC locus. To this end, we imputed the classical *HLA* alleles in BBJ and UKB, and performed association tests for 159 disease endpoints and 38 biomarkers (**Figure 3a** and **3b**). After the fine-mapping and conditional analyses (see **Methods**), we identified 75 and 129 independent association signals in BBJ and UKB, respectively (*P*<5.0×10^-8^; **Supplementary Table 8**). Among 53 and 63 traits associated with MHC in BBJ and UKB, 2 and 9 traits had never been previously shown to be associated with MHC, respectively. Overall, *HLA-B* in class I and *HLA-DRB1* in class II harbored the largest number of associations in both BBJ and UKB. For example, we fine-mapped the strong signal associated with PTB to HLA-DRβ1 Ser57 (OR=1.20, *P*=7.1×10^-19^) in BBJ. This is the third line of evidence showing the robust association of HLA with tuberculosis identified to date^28, 29^, and we newly fine-mapped the signal to *HLA-DRB1*. Interestingly, HLA-DRβ1 at position 57 also showed pleiotropic associations with other autoimmune and thyroid-related diseases, such as Grave’s disease (GD), hyperthyroidism, Hashimoto’s disease, Sjögren’s syndrome, chronic hepatitis B, and atopic dermatitis in BBJ. Of note, the effect direction of the association of HLA-DRβ1 Ser57 was the same between hyperthyroid status (OR=1.29, *P*=2.6×10^-14^ in GD and OR=1.37, *P*=1.4×10^-8^ in hyperthyroidism) and hypothyroid status (OR=1.50, *P*=9.0×10^-8^ in Hashimoto’s disease and OR=1.31), despite the opposite direction of thyroid hormone abnormality. This association of HLA-DRβ1 was also observed in Sjogren’s syndrome (OR=2.04, *P*=7.9×10^-12^), which might underlie epidemiological comorbidities of these diseases^30^. Other novel associations in BBJ included HLA-DRβ1 Asn197 with sarcoidosis (OR=2.07, *P*=3.7×10^-8^), and four independent signals with chronic sinusitis (i.e., *HLA-DRA*, *HLA-B*, *HLA-A*, and *HLA-DQA1*).

**Figure 3.**
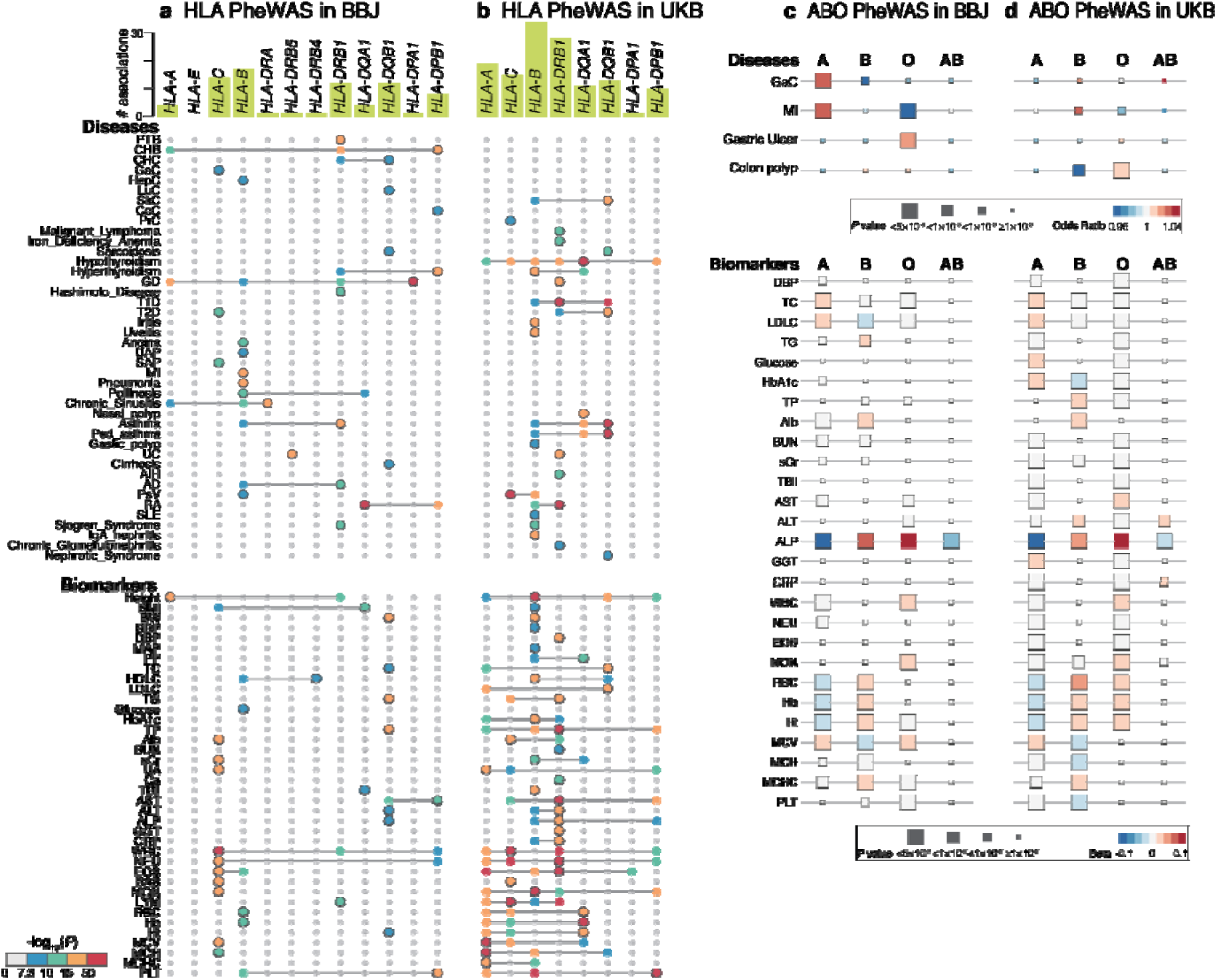
HLA and ABO association PheWAS. (**a**,**b**) Significantly associated HLA genes identified by HLA PheWAS in BBJ (**a**) or in UKB (**b**) are plotted (*P*<5.0×10^-8^). In addition to primary association signals of the phenotypes, independent associations identified by conditional analyses are also plotted, and the primary association is indicated by the plots with a gray border. The color of each plot indicates two-tailed *P* values calculated with logistic regression (for binary traits) or linear regression (for quantitative traits) as designated in the color bar at the bottom. The bars in green at the top indicate the number of significant associations per gene in each of the populations. The detailed allelic or amino acid position as well as statistics in the association are provided in **Supplementary Table 8**. (**c**,**d**) Significant associations identified by ABO blood-type PheWAS in BBJ (**c**) or in UKB (**d**) are shown as boxes and colored based on the odds ratio. The size of each box indicates two-tailed *P* values calculated with logistic regression (for binary traits) or linear regression (for quantitative traits).

Another representative pleiotropic locus in the human genome is the *ABO* locus. We performed ABO blood-type PheWAS in BBJ and UKB (**Figure 3c** and **3d**). We estimated the ABO blood type from three variants (rs8176747, rs8176746, and rs8176719 at 9q34.2)^31^, and associated them with the risk of diseases and quantitative traits for each blood group. A variety of phenotypes, including common diseases such as myocardial infarction as well as biomarkers such as blood cell traits and lipids, were associated with the blood types in both biobanks (**Supplementary Table 9**). We replicated an increased risk of gastric cancer in blood-type A as well as an increased risk of gastric ulcer in blood-type O in BBJ^32^.

### Genetic correlation elucidates the shared phenotypic domains across populations

The interplay between polygenicity and pleiotropy suggests widespread genetic correlations among complex human traits^33^. Genetic relationships among human diseases have contributed to the refinement of disease classifications^34^ and elucidation of the biology underlying the epidemiological comorbidity^33^. To obtain insights into the interconnections among human traits and compare them across populations, we computed pairwise genetic correlations (*r*_g_) across 106 traits (in Japanese) and 148 traits (in Europeans) with Z-score for *h*^2^ >2, using bivariate LD score regression (see **Methods**). We then defined the correlated trait domains by greedily searching for the phenotype blocks with pairwise *r*_g_>0.7 within 70% of *r*_g_ values in the block on the hierarchically clustered matrix of pairwise *r*_g_ values (**Extended Data Figure 11**). We detected domains of tightly correlated phenotypes, such as (i) cardiovascular-acting medications, (ii) coronary artery disease, (iii) type 2 diabetes-related phenotypes, (iv) allergy-related phenotypes, and (v) blood-cell phenotypes in BBJ (**Extended Data Figure 11a**). These domains implicated the shared genetic backgrounds on the similar diseases and their treatments (e.g., (ii) diseases of the circulatory system in ICD10 and their treatments) and diagnostic biomarkers (e.g., (iii) glucose and HbA1c in type 2 diabetes). Intriguingly, the corresponding trait domains were mostly identified in UKB as well (**Extended Data Figure 11b**). We considered that the current clinical boundaries for human diseases might broadly reflect the shared genetic etiology across populations, despite differences in populations and despite potential differences in diagnostic, environmental, and prescription practices.

### Deconvolution of a matrix of summary statistics of 159 diseases provides novel insights into disease pathogenesis

A major challenge in genetic correlation is that the *r*_g_ is a scalar value between two traits, which collapses the correlation over the whole genome into an averaged metric^35^. This approach is not straightforward in specifying a set of genetic variants driving the observed correlation, which would pinpoint biological pathways explaining the shared pathogenesis. To address this, genetic association statistics of diverse phenotypes can implicate latent structures underlying genotype-phenotype associations without a prior hypothesis. In particular, matrix decomposition of GWAS statistics is a promising approach^8, 36, 37^, which derives orthogonal components that explain association variance across multiple traits. This decomposition addresses two challenges in current genetic correlation studies. First, it informs us of genetic variants that explain the shared structure across multiple diseases, thereby enabling functional interpretation of the component. Second, it can be applicable to sub-significant associations, which are important in understanding contribution of common variants in rare diseases^36^ or in genetic studies in underrepresented populations where lower statistical power is inevitable.

Therefore, we applied DeGAs^8^ on a matrix of the disease GWAS summary statistics in Japanese and Europeans (*n*_disease_=159; **Figure 4a** and **4b**). To interpret the derived latent components, we annotated the genetic variants explaining each component (i) through GREAT genomic region ontology enrichment analysis^38^, (ii) through identification of relevant cell types using tissue specific regulatory DNA (ENCODE3^39^) and expression (GTEx^40^) profiles, and (iii) by projecting biomarker GWASs in BBJ and UKB (*n*_biomarker_=38) or metabolome GWASs in EAS cohort of Tohoku Medical Megabank Organization (ToMMo; **Methods**) and EUR cohort^41^ (*n*_metabolite_EAS_=206, *n*_metabolite_EUR_=248) into the component space derived from disease genetics (**Figure 4a**). We applied TSVD on the sparse Z score matrix of 22,980 variants, 159 phenotypes each in 2 populations (Japanese and Europeans), and derived 40 components that together explained 36.7% of the variance (**Extended Data Figure 12, 13**).

**Figure 4.**
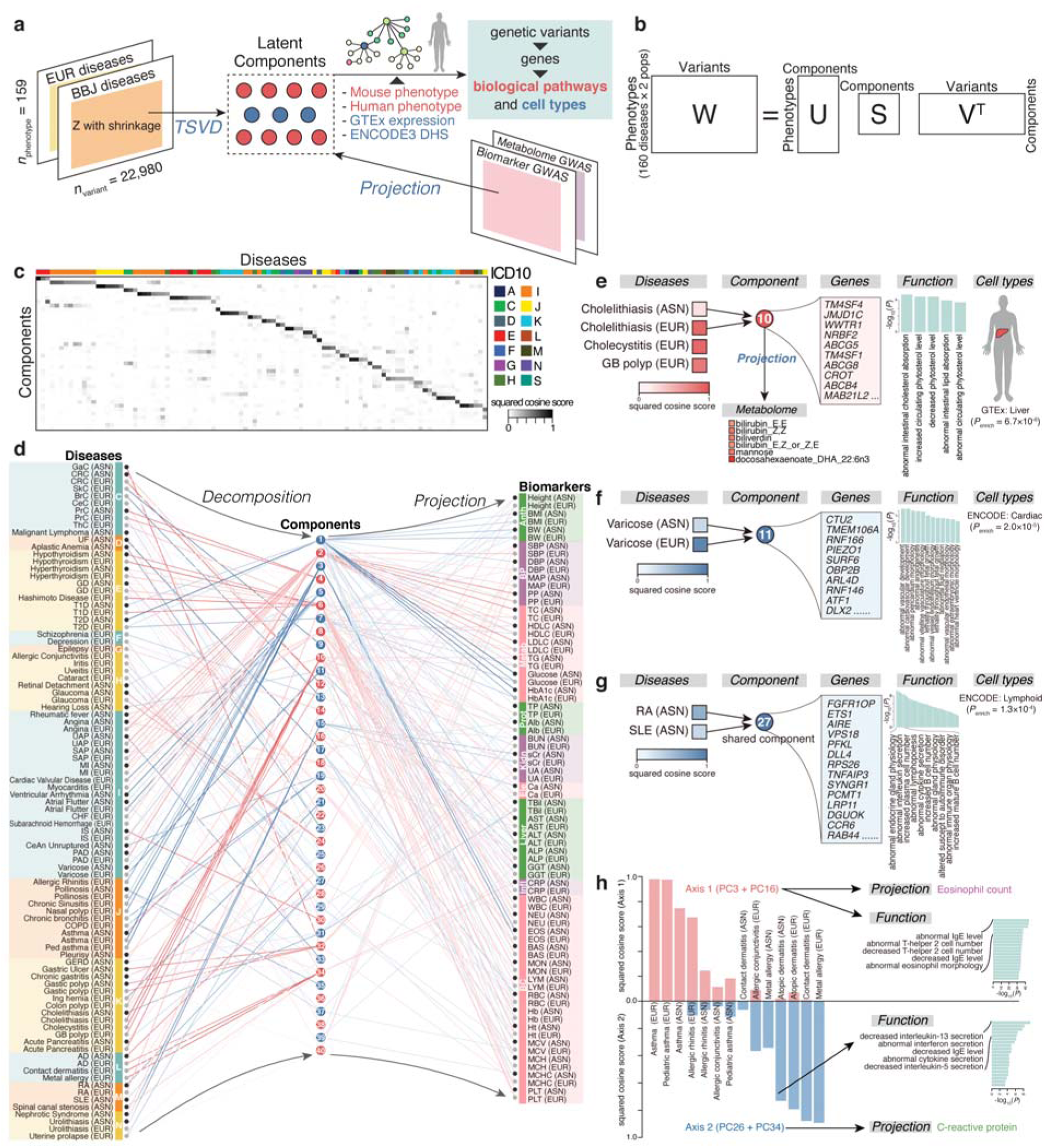
The deconvolution analysis of a matrix of summary statistics of 159 diseases across populations. (**a**) An illustrative overview of deconvolution-projection analysis. Using DeGAs framework, a matrix of summary statistics from two populations (EUR: European and BBJ: Biobank Japan) was decomposed into latent components, which were interpreted by annotation of a set of genetic variants driving each component and in the context of other GWASs through projection. (**b**) A schematic representation of TSVD applied to decompose a summary statistic matrix **W** to derive latent components. **U**, **S**, and **V** represent resulting matrices of singular values (**S**) and singular vectors (**U** and **V**). (**c**) A heatmap representation of DeGAs squared cosine scores of diseases (columns) to components (rows). The components are shown from 1 (top) to 40 (bottom), and diseases are sorted based on the contribution of each component to the disease measured by the squared cosine score (from component 1 to 40). Full results with disease and component labels are in **Extended Data Figure 15**. (**d**) Results of TSVD of disease genetics matrix and the projection of biomarker genetics. Diseases (left) and biomarkers (right) are colored based on the ICD10 classification and functional categorization, respectively. The derived components (middle; from 1 to 40) are colored alternately in blue or red. The squared cosine score of each disease to each component and each biomarker to each component is shown as red and blue lines. The width of the lines indicates the degree of contribution. The diseases with squared cosine score>0.3 in at least one component are displayed. Anth; anthropometry, BP; blood pressure, Metab; metabolic, Prot; protein, Kidn; kidney-related, Ele; Electrolytes, Liver; liver-related, Infl; Inflammatory, BC; blood cell. (**e**-**h**) Examples of disease-component correspondence and biological interpretation of the components by projection and enrichment analysis using GREAT. A representative component explaining a group of diseases based on the contribution score, along with responsible genes, functional enrichment results GREAT, relevant tissues, and relevant biobarkers/metabolites is shown. GB; gallbladder. RA; rheumatoid arthritis. SLE; systemic lupus erythematosus.

Globally, similar diseases as defined by the conventional ICD10 classification were explained by the same components, based on DeGAs trait squared cosine scores that quantifies component loadings^8^ (**Figure 4c, d**). This would be considered as a hypothesis-free support of the historically defined disease classifications. For example, component 1 explained the genetic association patterns of diabetes (E10 and E11 in ICD10) and component 2 explained those of cardiac and vascular diseases (I00-I83), in both populations. Functional annotation of the genetic variants explaining these components showed that component 1 (diabetes component) was associated with abnormal pancreas size (binomial *P*_enrichment_ =7.7×10^-19^) as a human phenotype, whereas component 2 (cardiovascular disease component) was associated with xanthelasma (i.e., cholesterol accumulation on the eyelids; binomial *P*_enrichment_ =3.0×10^-10^). Further, the genes comprising component 1 were enriched in genes specifically expressed in the pancreas (*P*_enrichment_ =5.5×10^-4^), and those comprising component 2 were enriched in genes specifically expressed in the aorta (*P*_enrichment_ =1.9×10^-3^; **Extended Data Figure 14**). By projecting the biomarker GWASs in BBJ and UKB and the metabolite GWASs in independent cohorts of EAS and EUR into this component space, we observed that component 1 represented the genetics of glucose and HbA1c, and component 2 represented the genetics of blood pressure and lipids, which are biologically relevant. This deconvolution-projection analyses suggested the latent genetic structure behind human diseases, which recapitulated the underlying biological functions, relevant tissues, and associated markers.

The latent components shared across diseases explained the convergent biology behind etiologically similar diseases. For example, component 10 explained the genetics of cholelithiasis (gall stone), cholecystitis (inflammation of gallbladder), and gall bladder polyp (**Figure 4e**). The projection of publicly available EUR metabolite GWASs into the component space identified that component 10 represented the bilirubin metabolism pathway. Component 10 was composed of variants involved in intestinal cholesterol absorption in mouse phenotype (binomial *P*_enrichment_=3.8×10^-10^). This is biologically relevant, since increased absorption of intestinal cholesterol is a major cause of cholelithiasis, which also causes cholecystitis^42^. The projection of the metabolite GWASs in independent Japanese cohort of ToMMo showed the connection between the component 10 and glycine that conjugates with bile acids^43^.

Some components were further utilized to interpret the underpowered GWAS with the use of well-powered GWAS, and to identify the contributor of shared genetics between different diseases. For example, we complemented underpowered varicose GWAS in BBJ (*n*_case_=474, genome-wide significant loci=0) with more powered GWAS in Europeans (*n*_case_=22,037, genome-wide significant loci=70). Both GWASs were mostly represented by component 11, which was explained by variants related to abnormal vascular development (binomial *P*_enrichment_=4.2×10^-7^; **Figure 4f**). Another example is component 27, which was shared with rheumatoid arthritis and systemic lupus erythematosus, two distinct but representative autoimmune diseases. Component 27 was explained by the variants associated with interleukin secretion and plasma cell number (binomial *P*_enrichment_=6.1×10^-10^ and 9.3×10^-10^, respectively), and significantly enriched in the DNase I hypersensitive site (DHS) signature of lymphoid tissue (*P*_enrichment_=1.3×10^-4^; **Figure 4g**). This might suggest the convergent etiology of the two autoimmune diseases, which was not elucidated by the genetic correlation alone.

Finally, we aimed at hypothesis-free categorization of diseases based on these components. Historically, hypersensitivity reactions have been classified into four types (e.g., types I to IV)^44^, but the clear sub-categorization of allergic diseases based on this pathogenesis and whether the categorization can be achieved solely by genetics were unknown. In TSVD results, the allergic diseases (mostly J and L in ICD10) were represented by the four components 3, 16, 26, and 34. By combining these components as axis-1 (components 3 and 16) and axis-2 (components 26 and 34), and comparing the cumulative variance explained by these axes, we defined *axis-1 dominant allergic diseases* (e.g., asthma and allergic rhinitis) and *axis-2 dominant allergic diseases* (metal allergy, contact dermatitis, and atopic dermatitis; **Figure 4h**). Intriguingly, the *axis-1 dominant diseases* etiologically corresponded to type I allergy (i.e., immediate hypersensitivity). The variants explaining *axis-1* were biologically related to IgE secretion and Th_2_ cells (binomial *P*_enrichment_=9.9×10^-46^ and 2.9×10^-44^, respectively). Furthermore, GWAS of eosinophil count was projected onto *axis-1*, which recapitulated the biology of type I allergy^45^. In contrast, the *axis-2 dominant diseases* corresponded to type IV allergy (i.e., cell-mediated delayed hypersensitivity). The variants explaining *axis-2* were associated with IL-13 and interferon secretion (binomial *P*_enrichment_=1.6×10^-10^ and 5.2×10^-9^, respectively), and GWAS of C-reactive protein was projected onto *axis-2*, which was distinct from *axis-1*^46^. To summarize, our deconvolution approach (i) recapitulated the existing disease classifications, (ii) implicated underlying biological mechanisms and relevant tissues shared among related diseases, and (iii) suggested potential application for genetics-driven categorization of human diseases.

## Discussion

Here, we performed 220 GWASs of human traits by incorporating the PMH and EMR data in BBJ, substantially expanding the atlas of genotype-phenotype associations in non-Europeans. We note that we additionally discovered 92 loci across 38 disease endpoints of which we had previously conducted GWASs in BBJ^12^, which highlights the value of curating PMH and EMR in biobanks. We then systematically compared their genetic basis with GWASs of corresponding phenotypes in Europeans. We confirmed the global replication of loci identified in BBJ, and discovered 5,343 novel loci through cross-population meta-analyses, highlighting the value of conducting GWASs in diverse populations. The results are openly shared through web resources, which will be a platform to accelerate further research such as functional follow-up studies and drug discovery^47^. Of note, leveraging these well-powered GWASs, we observed that the genes associated with endocrine/metabolic, circulatory, and respiratory diseases (E, I, and J by ICD10) were systematically enriched in targets of approved medications treating those diseases^48^ (**Extended Data Figure 16**). This should motivate us to use this expanded resource for genetics-driven novel drug discovery and drug repositioning.

The landscape of regional pleiotropy was globally shared across populations, and pleiotropic regions tended to have been under recent positive selection. One limitation of the current analysis is that we did not conduct statistical fine-mapping for every locus we identified, which might cause a concern over potential effects due to LD tagging for observed pleiotropy. Although we confirmed that the same pleiotropic variants were included within the 95% credible set for representative loci (*ALDH2* and *GCKR*; see **Supplementary Note**), more comprehensive statistical fine-mapping would further illuminate a global landscape of pleiotropic variants in the future. Moreover, elucidation of pleiotropy in other populations is warranted to replicate our results. To highlight the utility of deep phenotype GWASs, we decomposed the cross-population genotype–phenotype association patterns by TSVD. The latent components derived from TSVD showed the convergent biological mechanisms and relevant cell types across diseases, which can be utilized for re-evaluation of existing disease classifications. The incorporation of biomarker and metabolome GWAS summary statistics enabled interpretation of the latent components. Our approach might suggest a potential avenue for restructuring of medical diagnoses through dissecting the shared genetic basis across a spectrum of diseases, as analogous to the current disease classifications historically and empirically shaped through categorization of key symptoms across a spectrum of organ dysfunctions. However, we note that one major challenge in the deconvolution analyses is that the derived components and their order are affected by selection of phenotypes analyzed as input matrices. Thus, external validations are necessary before being generalized and used for refinement of disease categorization.

In conclusion, our study substantially expanded the atlas of genetic associations, supported the historically-defined categories of human diseases, and should accelerate the discovery of the biological basis contributing to complex human diseases.

## Data Availability

The genotype data of BBJ used in this study are available from the Japanese Genotype-phenotype Archive (JGA; http://trace.ddbj.nig.ac.jp/jga/index_e.html) with accession code JGAD00000000123 and JGAS00000000114. The UKB analysis was conducted via application number 47821. This study used the FinnGen release 3 data. Summary statistics of BBJ GWAS and trans-ethnic meta-analysis will be publicly available without any restrictions.

https://pheweb.jp/

## Acknowledgments

We sincerely thank all the participants of BioBank Japan, UK Biobank, and FinnGen. We thank Dr. Kyoko Watanabe for her input in the analysis of phenotypic correlations and pleiotropy. This research was supported by the Tailor-Made Medical Treatment program (the BioBank Japan Project) of the Ministry of Education, Culture, Sports, Science, and Technology (MEXT), the Japan Agency for Medical Research and Development (AMED). The FinnGen project is funded by two grants from Business Finland (HUS 4685/31/2016 and UH 4386/31/2016) and nine industry partners (AbbVie, AstraZeneca, Biogen, Celgene, Genentech, GSK, MSD, Pfizer and Sanofi). Following biobanks are acknowledged for collecting the FinnGen project samples: Auria Biobank (https://www.auria.fi/biopankki/), THL Biobank (https://thl.fi/fi/web/thl-biopank), Helsinki Biobank (https://www.terveyskyla.fi/helsinginbiopankki/), Northern Finland Biobank Borealis (https://www.ppshp.fi/Tutkimus-ja-opetus/Biopankki), Finnish Clinical Biobank Tampere (https://www.tays.fi/biopankki), Biobank of Eastern Finland (https://ita-suomenbiopankki.fi), Central Finland Biobank (https://www.ksshp.fi/fi-FI/Potilaalle/Biopankki), Finnish Red Cross Blood Service Biobank (https://www.bloodservice.fi/Research%20Projects/biobanking), Terveystalo Biobank Finland (https://www.terveystalo.com/fi/Yritystietoa/Terveystalo-Biopankki/Biopankki/). S.S. was in part supported by The Mochida Memorial Foundation for Medical and Pharmaceutical Research, Kanae Foundation for the promotion of medical science, Astellas Foundation for Research on Metabolic Disorder, and The JCR Grant for Promoting Basic Rheumatology M.Kanai was supported by a Nakajima Foundation Fellowship and the Masason Foundation. Y.Tanigawa is in part supported by a Funai Overseas Scholarship from the Funai Foundation for Information Technology and the Stanford University School of Medicine. M.A.R. is in part supported by National Human Genome Research Institute (NHGRI) of the National Institutes of Health (NIH) under award R01HG010140 (M.A.R.), and a National Institute of Health center for Multi- and Cross-population Mapping of Mendelian and Complex Diseases grant (5U01 HG009080). The content is solely the responsibility of the authors and does not necessarily represent the official views of the National Institutes of Health. Y.O. was supported by the Japan Society for the Promotion of Science (JSPS) KAKENHI (19H01021, 20K21834), and AMED (JP20km0405211, JP20ek0109413, JP20ek0410075, JP20gm4010006, and JP20km0405217), Takeda Science Foundation, and Bioinformatics Initiative of Osaka University Graduate School of Medicine, Osaka University.

## Author Contributions

S.S., M. Kanai, and Y.O. conceived the study. S.S., M. Kanai, Y. Tanigawa., M.A.R., and Y.O. wrote the manuscript. S.S., M. Kanai, J.K., M. Kurki, T.Konuma, Kenichi Yamamoto, M.A., K.Ishigaki, Kazuhiko Yamamoto, Y. Kamatani, A.P., M.J.D., and Y.O. conducted GWAS data studies. S.S., Y. Tanigawa., and M.A.R. conducted statistical decomposition analysis. S.S., S.T., A.N., G.T., and Y.O. conducted metabolome analysis. A.S., K.S., W.O., Ken Yamaji, K.T., S.A., Y.Takahashi, T.S., N.S., H.Y., S.Minami, S.Murayama, Kozo Yoshimori, S.N., D.O., M.H., A.M., Y.Koretsune, K.Ito, C.T., T.Y., I.K., T.Kadowaki, M.Y., Y.N., M.Kubo, Y.M., Kazuhiko Yamamoto, and K.M. collected and managed samples and data. A.P. and M.J.D. coordinated collaboration with FinnGen.

## Competing Financial Interests

M.A.R. is on the SAB of 54Gene and Computational Advisory Board for Goldfinch Bio and has advised BioMarin, Third Rock Ventures, MazeTx and Related Sciences. The funders had no role in study design, data collection and analysis, decision to publish, or preparation of the manuscript.

## Data availability

The genotype data of BBJ used in this study are available from the Japanese Genotype-phenotype Archive (JGA; http://trace.ddbj.nig.ac.jp/jga/index_e.html) with accession code JGAD00000000123 and JGAS00000000114. The UKB analysis was conducted via application number 47821. This study used the FinnGen release 3 data. All summary statistics of 220 GWASs (BioBank Japan, European, and cross-population meta-analyses) are deposited at the National Bioscience Database Center (NBDC) Human Database with the accession code hum0197. We also provide an interactive visualization of Manhattan, Locus Zoom, and PheWAS plots with downloadable GWAS summary statistics at our PheWeb.jp website [https://pheweb.jp/]. The summary statistics of metabolite GWASs in the Japanese population (Tohoku Medical Megabank Organization) are being prepared as a different project. The previous version of the partial statistics is publicly available at https://jmorp.megabank.tohoku.ac.jp/202008/gwas/TGA000003.

## Code availability

We used publicly available software for the analyses. The used software is listed and described in the **Method** section of our manuscript.

## Methods

### Genome-wide association study of 220 traits in BBJ

We conducted 220 deep phenotype GWASs in BBJ. BBJ is a prospective biobank that collaboratively collected DNA and serum samples from 12 medical institutions in Japan and recruited approximately 200,000 participants, mainly of Japanese ancestry (**Supplementary Note**). All study participants had been diagnosed with one or more of 47 target diseases by physicians at the cooperating hospitals. We previously conducted GWASs of 42 out of the 47 target diseases^12^. In this study, we newly curated the PMH records included in the clinical data, and performed text-mining to retrieve disease records from the free-format EMR as well. For disease phenotyping, PMH record has already been curated and formatted as sample × phenotype table. Regarding EMR, we searched for Japanese term of a given disease diagnosis in the cells designated as the presence of PMH, which was compiled into sample × phenotype table. We merged both information with the target disease status, and defined the case status for 159 diseases with a case count>50 (**Supplementary Table 2**). As controls, we used samples in the cohort without a given diagnosis or related diagnoses, which was systematically defined by using the phecode framework^3^ (**Supplementary Table 1**). For medication-usage phenotyping, we again retrieved information by text-mining of 7,018,972 medication records. Then, we categorized each medication trade name by using the ATC (Anatomical Therapeutic Chemical Classification), World Health Organization, which is used for the classification of active ingredients of drugs according to the organ or system on which they act and their therapeutic, pharmacological and chemical properties. For biomarker phenotyping, we used the same processing and quality control method as previously described (**Supplementary Table 2** for phenotype summary)^13, 49^. In brief, we generally used the laboratory values measured at the participants’ first visit to the recruitment center, and excluded measurements outside three times of interquartile range (IQR) of upper/lower quartile across participants. For individuals taking anti-hypertensive medications, we added 15 mmHg to systolic blood pressure (SBP) and 10 mmHg to diastolic blood pressure (DBP). For individuals taking a statin, we applied the following correction to the lipid measurements: i) Total cholesterol was divided by 0.8; ii) measured LDL-cholesterol (LDLC) was adjusted as LDLC / 0.7; iii) derived LDLC from the Friedewald was re-derived as (Total cholesterol / 0.8) - HDLC - (Triglyceride/ 5).

We genotyped participants with the Illumina HumanOmniExpressExome BeadChip or a combination of the Illumina HumanOmniExpress and HumanExome BeadChips. Quality control of participants and genotypes was performed as described elsewhere^14^. In this project, we analyzed 178,726 participants of East Asian ancestry as estimated by the principal component analysis (PCA)-based sample selection criteria. The genotype data were further imputed with 1000 Genomes Project Phase 3 version 5 genotype (*n*=2,504) and Japanese whole-genome sequencing data (*n*=1,037) using Minimac3 software. After this imputation, we excluded variants with an imputation quality of Rsq<0.7, resulting in 13,530,797 variants analyzed in total.

We conducted GWASs for binary traits (i.e., disease endpoints and medication usage) by using a generalized linear mixed model implemented in SAIGE (version 0.37), which had substantial advantages in terms of (i) maximizing the sample size by including genetically related participants, and (ii) controlling for case–control imbalance^15^, which was the case in many of the disease endpoints in this study. We included adjustments for age, age^2^, sex, age×sex, age^2^×sex, and top 20 principal components for as covariates used in step 1. For sex-specific diseases, we alternatively adjusted for age, age^2^, and the top 20 principal components as covariates used in step 1, and we used only controls of the sex to which the disease is specific. We conducted GWASs for quantitative traits (i.e., biomarkers) by using a linear mixed model implemented in BOLT-LMM (version 2.3.4). We included the same covariates as used in the binary traits above.

All the participants provided written informed consent approved from ethics committees of the Institute of Medical Sciences, the University of Tokyo and RIKEN Center for Integrative Medical Sciences.

### Harmonized genome-wide association study of 220 traits in UKB and FinnGen

We conducted the GWASs harmonized with BBJ in UKB and in FinnGen. The UK Biobank project is a population-based prospective cohort that recruited approximately 500,000 people across the United Kingdom (**Supplementary Note**). We defined case and control status of 158 disease endpoints, which were originally retrieved from the clinical information in UKB and mapped to BBJ phenotypes via phecode (**Supplementary Table 1**). We also analyzed 38 biomarker values provided by the UKB. The genotyping was performed using either the Applied Biosystems UK BiLEVE Axiom Array or the Applied Biosystems UK Biobank Axiom Array. The genotypes were further imputed using a combination of the Haplotype Reference Consortium, UK10K, and 1000 Genomes Phase 3 reference panels by IMPUTE4 software^10^. In this study, we analyzed 361,194 individuals of white British genetic ancestry as determined by the PCA-based sample selection criteria (see **URLs**). We excluded the variants with (i) INFO score ≤ 0.8, (ii) MAF ≤ 0.0001 (except for missense and protein-truncating variants annotated by VEP^50^, which were excluded if MAF ≤ 1 × 10^-6^), and (iii) *P*_HWE_ ≤ 1 × 10, resulting in 13,791,467 variants analyzed in total. We conducted GWASs for 159 disease endpoints by using SAIGE with the same covariates used in the BBJ GWAS. For biomarker GWASs, we used publicly available summary statistics of UKB biomarker GWAS when available (see **URLs**), and otherwise performed linear regression using PLINK software with the same covariates, excluding the genetically related individuals (the 1st, 2nd, or 3rd degree)^10^. For medication usage GWASs, we used publicly available summary statistics of medication usage in UKB^20^, which was organized by the ATC and thus could be harmonized with BBJ GWASs.

FinnGen is a public–private partnership project combining genotype data from Finnish biobanks and digital health record data from Finnish health registries (**Supplementary Notes**). For GWASs, we used the summary statistics of FinnGen release 3 data (see **URLs**). The disease endpoints were mapped to BBJ phenotypes by using ICD10 code, and we defined 128 out of 159 endpoints in BBJ. The genome coordinates in summary statistics were lifted over to hg19, and we analyzed 16,859,359 variants after QC. We did not conduct biomarker and medication-related GWASs because the availability of these phenotypes was limited.

### Meta-analysis, definition of significant loci, and annotation of the lead variants with genome-wide significance

First, we performed intra-European meta-analysis when summary statistics of both UKB and FinnGen were available, and then performed cross-population meta-analysis across three or two cohorts in 159 disease endpoints, 38 biomarker values, and 23 medication usage GWASs. We conducted these meta-analyses by using the inverse-variance method and estimated heterogeneity with Cochran’s Q test with METAL software^51^. In this meta-analysis, we included all variants after QC in each of the three cohorts. The overlapping variants among the cohorts are summarized in **Extended Data Figure 17**. The summary statistics of primary GWASs in BBJ and cross-population meta-analysis GWASs are openly shared without any restrictions.

We adopted the conventional genome-wide significance threshold of<5.0×10^-8^, as well as considering the Bonferroni-corrected threshold of<7.6×10^-11^ (5.0×10^-8^ / (220 phenotypes × 3 populations)) in the context of cross-population meta-analysis. We defined independent genome-wide significant loci on the basis of genomic positions within ±500 kb from the lead variant. We considered a trait-associated locus as novel when the locus within ±1 Mb from the lead variant did not include any variants that were previously reported to be significantly associated with the same disease.

To systematically collect previously reported significant associations (5.0×10^-8^) as known variants, we

1. exhaustively searched for previous reports of genetic association in a given trait using the GWAS Catalog^4^, since it is currently recognized as a standard and most comprehensive database of genetic associations,
2. systematically searched PubMed when the corresponding trait was not included in GWAS Catalog, and
3. exceptionally included preprints in case we have collaboratively worked on them, in order to avoid duplicated publication.

The goal was to comprehensively include only robust and invariant associations. In this way, we included 75,230 associations across 181 traits in 1,792 literatures as of December 31, 2020 (**Supplementary Table 10**).

We annotated the lead variants using ANNOVAR software, such as rsIDs in dbSNP database (see **URLs**), the genomic region and closest genes, and functional consequences. We also supplemented this with the gnomAD database^18^, and also looked for the allele frequencies in global populations as an independent resource.

### Replication of significant associations in BBJ

For 2,287 lead variants in the genome-wide significant loci of 159 disease endpoints and 38 biomarkers in BBJ, we compared the effect sizes and directions with European-only meta-analysis when available and with UKB-based summary statistics otherwise. Of them, 1,929 variants could be compared with the corresponding European GWASs. Thus, we performed the Pearson’s correlation test for these variants’ beta in the association test in BBJ and in European GWAS. We also performed the correlation tests with variants with *P*_EUR_<0.05 and to those with *P*_EUR_<5.0×10^-8^.

### Cross-population genetic correlation

To estimate cross-population genetic-effect correlations between BBJ and European GWASs considering polygenic signals, we used Popcorn software (version 1.0)^52^. For this analysis, we restricted the traits to those with (i) heritability Z score from LDSC>2 (which will be explained later in **Methods**), and (ii) both BBJ and European heritability calculated by Popcorn>0.01. We excluded the MHC region from the analysis because of its complex LD structure. Using these QCed traits’ summary statistics, we calculated the cross-population genetic-effect correlation between EUR and EAS with precomputed cross-population scores for EUR and EAS 1000 Genomes populations provided by the authors.

### Evaluation of regional pleiotropy

We assessed the regional pleiotropy based on each tested genetic variant separately for BBJ GWASs and for European GWASs (i.e., intra-European meta-analysis when FinnGen GWAS was available and UKB summary statistics otherwise). We quantified the degree of pleiotropy per genetic variant by aggregating and counting the number of genome-wide significant associations across 220 traits. We then annotated loci from the largest number of associations (*n*_associations_ ≥ 9 in BBJ and ≥ 18 in Europeans) in **Figure 2a, b**.

Next, we assessed the recent natural selection signature within the pleiotropic loci separately for Japanese and for Europeans. To do this, we first defined the pleiotropic loci by identifying genetic variants that harbored a larger number of significant associations than a given threshold. We varied this threshold from 1 to 40. Then, at each threshold, we calculated the sum of SDS χ^2^ values within the pleiotropic loci, and compared this with the χ^2^ distribution under the null hypothesis with a degree of freedom equal to the number of variants in the loci. We thus estimated the SDS enrichment within the pleiotropic loci defined by a given threshold as fold change and *P* value. The SDS values were obtained from the web resource indicated in the original article on Europeans (see **URLs**) and provided by the authors on Japanese^27^. The raw SDS values were normalized according to the derived allele frequency as described previously.

### Fine-mapping of HLA and ABO loci

We performed the fine-mapping of MHC associations in BBJ and UKB by HLA imputation^53^. In BBJ, we imputed classical HLA alleles and corresponding amino acid sequences using the reference panel recently constructed from 1,120 individuals of Japanese ancestry by the combination of SNP2HLA software, Eagle, and minimac3, as described previously^54^. We applied post-imputation quality control to keep the imputed variants with minor allele frequency (MAF) ≥ 0.5% and Rsq>0.7. For each marker dosage that indicated the presence or absence of an investigated HLA allele or an amino acid sequence, we performed an association test with the disease endpoints and biomarkers. We assumed additive effects of the allele dosages on phenotypes in the regression models. We included the same covariates as in the GWAS. In UKB, we imputed classical HLA alleles and corresponding amino acid sequences using the T1DGC reference panel of European ancestry (*n*=5,225)^55^. We applied the same post-imputation quality control and performed the association tests as in BBJ.

### Heritability and genetic correlation estimation

We performed LD score regression (see **URLs**) for GWASs of BBJ and Europeans to estimate SNP-based heritability, potential bias, and pairwise genetic correlations. Variants in the MHC region (chromosome 6:25–34 Mb) were excluded. We also excluded variants with χ^2^ > 80, as recommended previously . For heritability estimation, we used the baselineLD model (version 2.2), which included 97 annotations that correct for bias in heritability estimates^57^. We note that we did not report liability-scale heritability, since population prevalence of 159 diseases in each country was not always available, and the main objective of this analysis was an assessment of bias in GWAS, rather than the accurate estimation of heritability. We calculated the heritability Z-score to assess the reliability of heritability estimation, and reported the LDSC results with Z-score for *h*^2^ is>2 (**Supplementary Table 3**). For calculating pairwise genetic correlation, we again restricted the target GWASs to those whose Z-score for *h*^2^ is>2, as recommended previously^56^. In total, we calculated genetic correlation for 106 GWASs in BBJ and 148 in European GWASs, which resulted in 5,565 and 10,878 trait pairs, respectively.

To illustrate trait-by-trait genetic correlation, we hierarchically clustered the *r*_g_ values with hclust and colored them as a heatmap (**Extended Data Figure 11**). To adopt reliable genetic correlations, we restricted the *r*_g_ values that had *P*_cor_<0.05. Otherwise, the *r*_g_ values were replaced with 0. We then defined the tightly clustered trait domains by greedily searching for the phenotype blocks with pairwise *r*_g_>0.7 within 70% of *r*_g_ values in the block from the top left of the clustered correlation matrix. We manually annotated each trait domain by extracting the characteristics of traits constituting the domain (**Extended Data Figure 11**).

### Deconvolution of a matrix of summary statistics by TSVD

We performed the TSVD on the matrix of genotype-phenotype association Z scores as described previously as DeGAs framework^8^. In this study, we first focused on 159 disease endpoint GWASs in BBJ and European GWAS (i.e., 318 in total) to derive latent components through TSVD. On constructing a Z-score matrix, we conducted variant-level QC. We removed variants located in the MHC region (chromosome 6: 25–34 Mb), and replaced unreliable Z-score estimates with zero when one of the following conditions were satisfied as in Tanigawa et al.^8^:

*P* value of marginal association ≥ 0.001
Standard error of beta value ≥ 0.2

Considering that rows and columns with all zeros do not contribute to matrix decomposition, we excluded variants that had all zero Z-scores across 159 traits in either in BBJ or Europeans. We then performed LD pruning using PLINK software^58^ (“--indep-pairwise 50 5 0.1”) with an LD reference of 5,000 randomly selected individuals of white British UKB participants to select LD-independent variant sets, which resulted in a total of 22,980 variants. Thus, we made a Z-score matrix (= **W**) with a size of 318 (*N*: 159 diseases × 2 populations) × 22,980 (*M*: variants). With a predetermined number of *K*, TSVD decomposed **W** into a product of three matrices: **U**, **S**, and **V^T^**: **W** = **USV^T^**. **U** = (*u_i,k_*)*_i,k_* is an orthonormal matrix of size *N* × *K* whose columns are phenotype singular vectors, **S** is a diagonal matrix of size *K*× *K* whose elements are singular values, and **V** = (*v_j,k_*)_j,k_ is an orthonormal matrix of size *M* × *K* whose columns are variant singular vectors. Here we set *K* as 40, which together explained 36.7% of the total variance of the original matrix. This value was determined by experimenting with different values from 20 to 100 and selecting the informative and sufficient threshold. We used the TruncatedSVD module in the sklearn.decomposition library of python for performing TSVD.

To interpret and visualize the results of TSVD, we calculated the squared cosine scores. The phenotype squared cosine score, 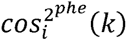, is a metric to quantify the relative importance of the *k*th latent component for a given phenotype *i*, and is defined as follows;

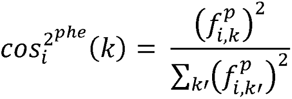

where

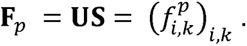

### Annotation of the components by using GREAT and identification of relevant cell types

We calculated the variant contribution score, which is a metric to quantify the contribution of a given variant *j* to a given component *k* as follows;

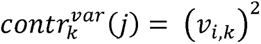

For each component, we can thus rank the variants based on their contribution to the component and calculate the cumulative contribution score. We defined a set of *contributing variants* to a given component to include top-ranked variants that had high contribution scores until the cumulative contribution score to the component exceeded 0.5. For these variant sets contributing to the latent components, we performed the GREAT (version 4.0.4) binomial genomic region enrichment analysis^38^ based on the size of the regulatory domain of genes and quantified the significance of enrichment in terms of binomial fold enrichment and binomial *P* value to biologically interpret these components. We used the human phenotype and mouse genome informatics phenotype ontology, which contains manually curated knowledge about the hierarchical structure of phenotypes and genotype-phenotype mapping of human and mouse, respectively. The enriched annotation with a false discovery rate (FDR)<0.05 is considered significant and displayed in the figures.

For a gene set associated with the contributing variants with a given component (*P*< 0.05), we sought to identify relevant cell types by integrating two datasets: (i) ENCODE3 DHS regulatory patterns across human tissues from non-negative matrix factorization (NFM)^39^ and (ii) specifically expressed genes defined from GTEx data^40^. In brief, a vocabulary (i.e., DHS patterns) for regulatory patterns was defined from the NFM of 3 million DHSs × 733 human biosamples encompassing 438 cell and tissue types. Then, for each regulatory vocabulary, GENCODE genes were assigned based on their overlying DHSs. The gene labeling result was downloaded from the journal website^39^. We also defined genes specifically expressed in 53 tissues from GTEx version 7 data, based on the top 5% of the *t*-statistics in each tissue as described elsewhere^59^. Then, for (i) each regulatory vocabulary and (ii) each tissue, we performed Fisher’s exact tests to investigate whether the genes associated with a given component are significantly enriched in the defined gene set.

### Projection of biomarker and metabolite GWASs into the component space

To further help interpret the latent components derived from disease-based TSVD, we projected the Z-score matrix of biomarker GWASs and metabolite GWASs into the component space. Briefly, we constructed the Z-score matrices (**W’**) of 38 biomarkers of BBJ and European GWASs (i.e., 76 rows) and 248 known metabolites of independent previous GWASs in the European population^41^ × 22,980 variants (**Supplementary Table 11**). Then, using the **V** from the disease-based TSVD, we calculated the phenotype contribution as follows;

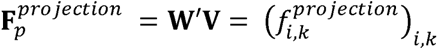

We note that for metabolite GWASs, since the GWASs were imputed with the HapMap reference panel, we imputed Z-scores of missing variants using ssimp software^60^ (version 0.5.5 --ref 1KG/EUR --impute.maf 0.01), and otherwise we set the missing Z-scores to zero.

### Projection of Metabolite GWASs in Japanese into the component space

To investigate whether the projection analysis is applicable to independent dataset, we conducted metabolite GWASs in ToMMo. ToMMo is a community-based biobank that combines medical and genome information from the participants in the Tohoku region of Japan^61^. Detailed cohort description is presented in **Supplementary Notes**. In this study, we analyzed a total of 206 metabolites^62^ measured by proton nuclear magnetic resonance (NMR) or liquid chromatography (LC)–MS (**Supplementary Table 12**). For sample QC, we excluded samples meeting any of the following criteria: (1) genotype call rate<95%, (2) one individual from each pair of those in close genetic relation (PI_HAT calculated by PLINK^58^ ≥ 0.1875) based on call rate, and (3) outliers from Japanese ancestry cluster based on the principal component analysis with samples of 1KGP phase 3 data. For phenotype QC, we excluded (1) the measurements in pregnant women, (2) those which took time from sampling to biobanking ≥ 2 days, and (3) phenotypic outlier defined as log-transformed measurements laying more than 4 SD from the mean for each metabolite. The participants were genotyped with a custom SNP array for the Japanese population (i.e., Japonica Array v2). For genotype QC, we excluded variants meeting any of the following criteria: (1) call rate<98%, (2) *P* value for Hardy–Weinberg equilibrium<1.0 × 10^−6^, and (3) MAF<0.01. The QCed genotype data were pre-phased by using SHAPEIT2 software (r837), and imputed by using IMPUTE4 software (r300.3) with a combined reference panel of 1KGP phase3 (*n*=2,504) and population specific WGS data (i.e., 3.5KJPNv2; *n*=3,552)^62^. After imputation, we excluded variants with imputation INFO<0.7. For GWASs, we obtained the residuals from a linear regression model of each of log-transformed metabolites adjusted for age, age^2^, sex, time period from sampling to biobanking, and top 20 genotype PCs. The residuals were then transformed by rank-based inverse normalization. Association analysis of imputed genotype dosage with the normalized residual of each metabolite was performed using PLINK2 software. We constructed the Z-score matrices (**W’**) of the Japanese metabolites GWASs (i.e., 206 rows) × 22,980 variants, in which we applied the same QC to the Z-scores and set the missing Z-scores to zero again. We then performed the projection as described above.

### Drug target enrichment analysis

To investigate whether disease-associated genes are systematically enriched in the targets of the approved drugs for the treatment of those diseases, the Genome for REPositioning drugs (GREP)^48^ was used. A list of genes closest to the lead variants from GWAS, which was concatenated based on the alphabetical category of ICD10 (A to N), was used as an input gene set to test the enrichment for the target genes of approved drugs for diseases of a given ICD10 category.

## URLs

- SDS values in UK10K provided by Pritchard’s lab; http://web.stanford.edu/group/pritchardlab/UK10K-SDS-values.zip
- Summary statistics of BBJ, European, and cross-population meta-analysis GWASs ; https://pheweb.jp/
- Summary statistics of biomarker GWASs in UKB by Neale’s lab ; http://www.nealelab.is/uk-biobank/ukbround2announcement
- LDSC software; https://github.com/bulik/ldsc
- FinnGen release 3 data; https://www.finngen.fi/en/access_results
- dbSNP; https://www.ncbi.nlm.nih.gov/snp/
- World Health Organization, Global Tuberculosis Report; https://www.who.int/tb/publications/global_report/en/
- Metabolite GWAS summary statistics in the European populations: http://metabolomics.helmholtz-muenchen.de/gwas/index.php?task=download

