## Supplementary Materials for "A global atlas of genetic associations of 220 deep phenotypes"

**Sakaue and Kanai et al.**

### Table of Contents:


Page 3 **Extended Data Figure 1**
Page 4 **Extended Data Figure 2**
Page 5 **Extended Data Figure 3**
Page 6 **Extended Data Figure 4**
Page 7 **Extended Data Figure 5**
Page 8 **Extended Data Figure 6**
Page 9 **Extended Data Figure 7**
Page 10 **Extended Data Figure 8**
Page 11 **Extended Data Figure 9**
Page 12 **Extended Data Figure 10**
Page 13 **Extended Data Figure 11**
Page 14 **Extended Data Figure 12**
Page 15 **Extended Data Figure 13**
Page 16 **Extended Data Figure 14**
Page 17 **Extended Data Figure 15**
Page 18 **Extended Data Figure 16**
Page 19 **Extended Data Figure 17**
Page 20 **Supplementary Notes**
Page 30 **Supplementary References**


**Supplementary Tables** are provided by a separate excel file.

**Extended Data Figure 1.**

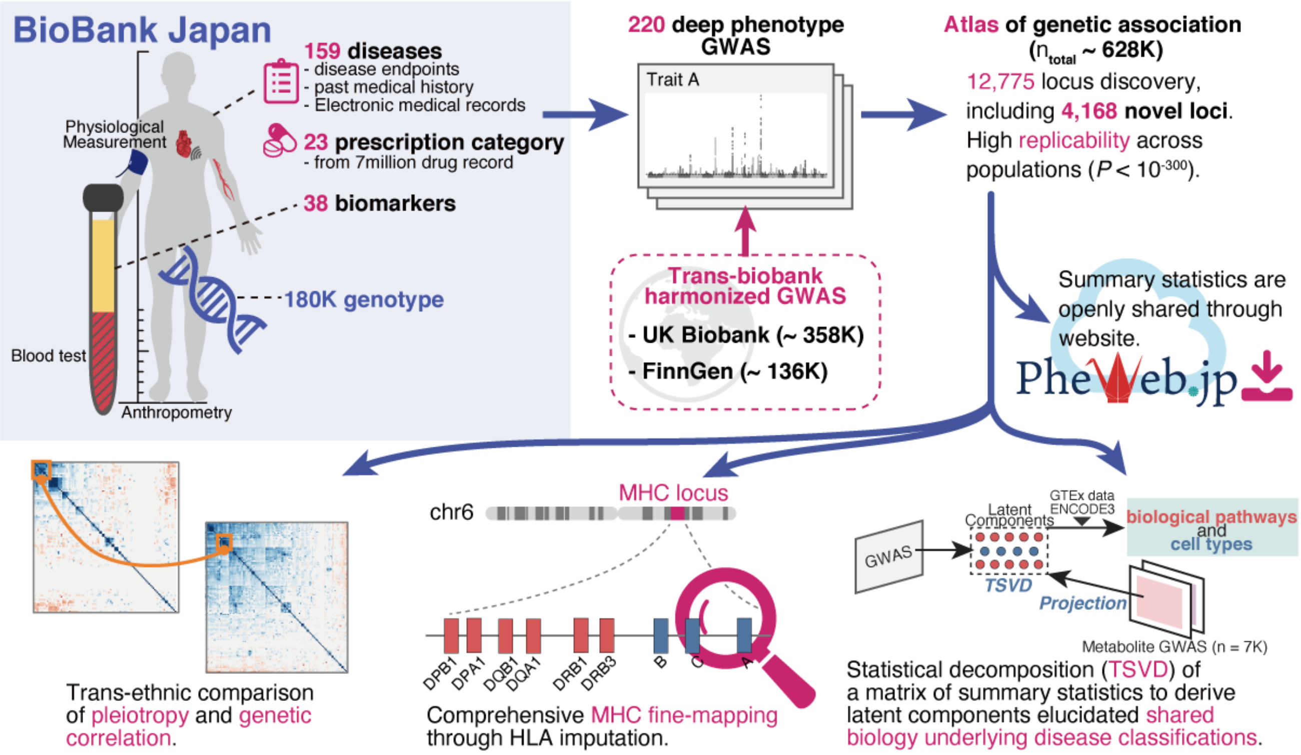


**Extended Data Figure 1 | Overview of this study.**

We performed 220 deep-phenotype GWASs in BioBank Japan, including 108 novel GWASs ever conducted in East Asian population. We performed trans-biobank meta-analyses with UK Biobank and FinnGen ($n_{total}$ = 628,000), resulting in discovery of 4,168 novel loci. All summary statistics will be openly shared through web database. As downstream analyses, we performed (i) cross-population comparison of pleiotropy and genetic correlation, (ii) comprehensive HLA fine-mapping, and (iii) statistical decomposition of a matrix of summary statistics to gain insights into biology underlying current disease classifications, by incorporating functional genomics, metabolomics, and biomarker data.

**Extended Data Figure 2.**

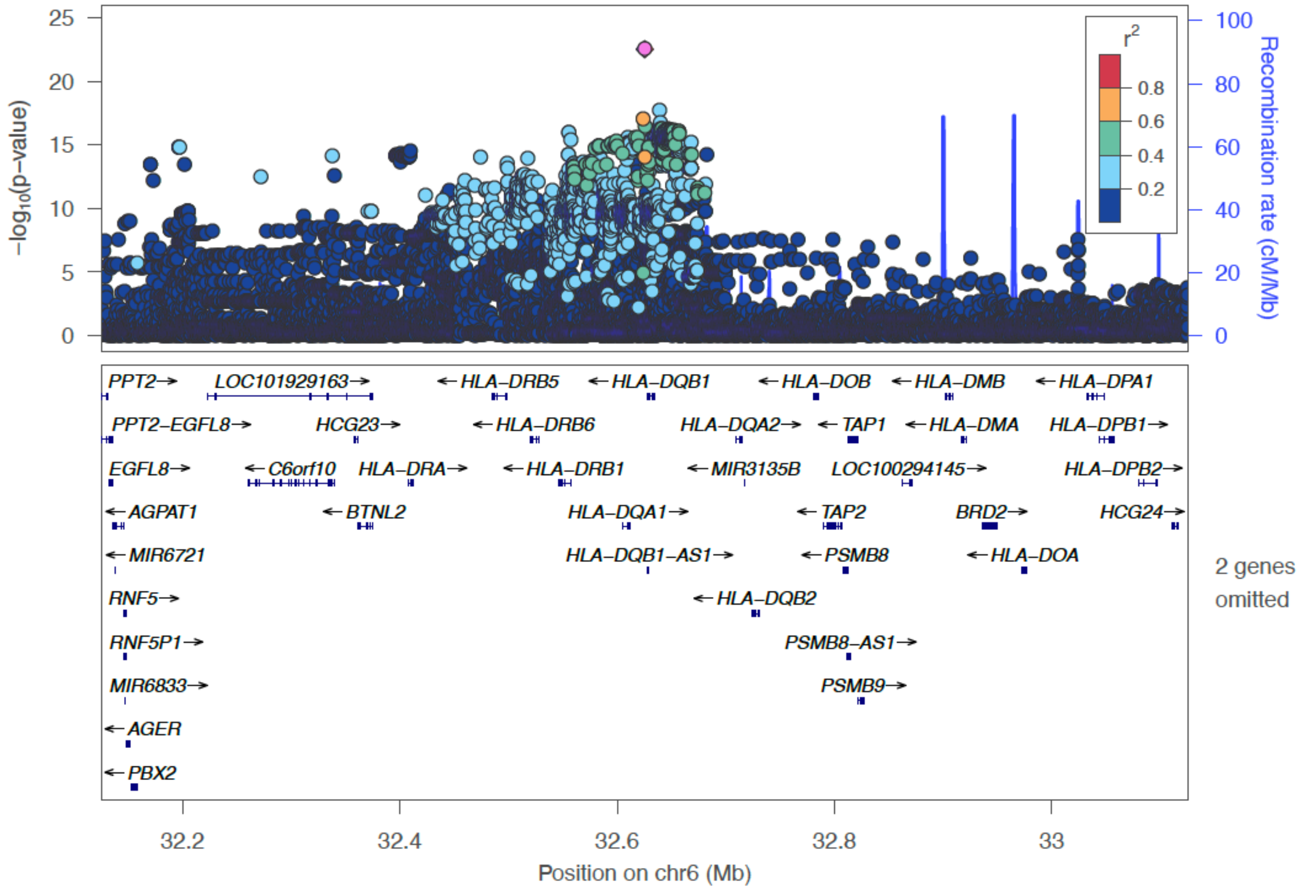


**Extended Data Figure 2 | Locus plot for Pulmonary Tuberculosis (PTB) in BBJ.**

Regional association plots are shown. The lead variant (rs140780894) is colored in pink, and colors of other dots indicate linkage disequilibrium measure $r^2$ with the lead variant.

**Extended Data Figure 3.**

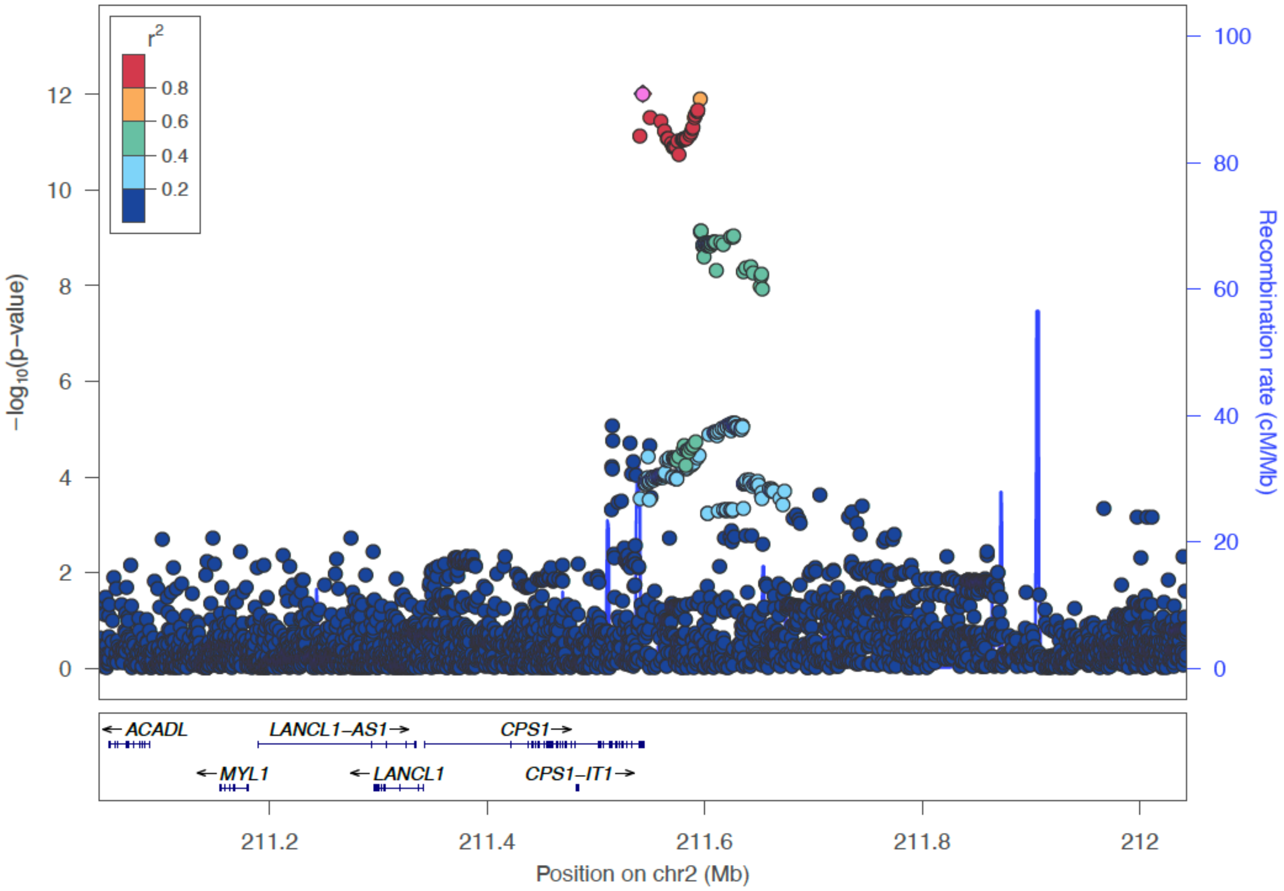


**Extended Data Figure 3 | Locus plot for cholelithiasis in BBJ.**

Regional association plots are shown. The lead variant (rs715) is colored in pink, and colors of other dots indicate linkage disequilibrium measure $r^2$ with the lead variant.

**Extended Data Figure 4.**

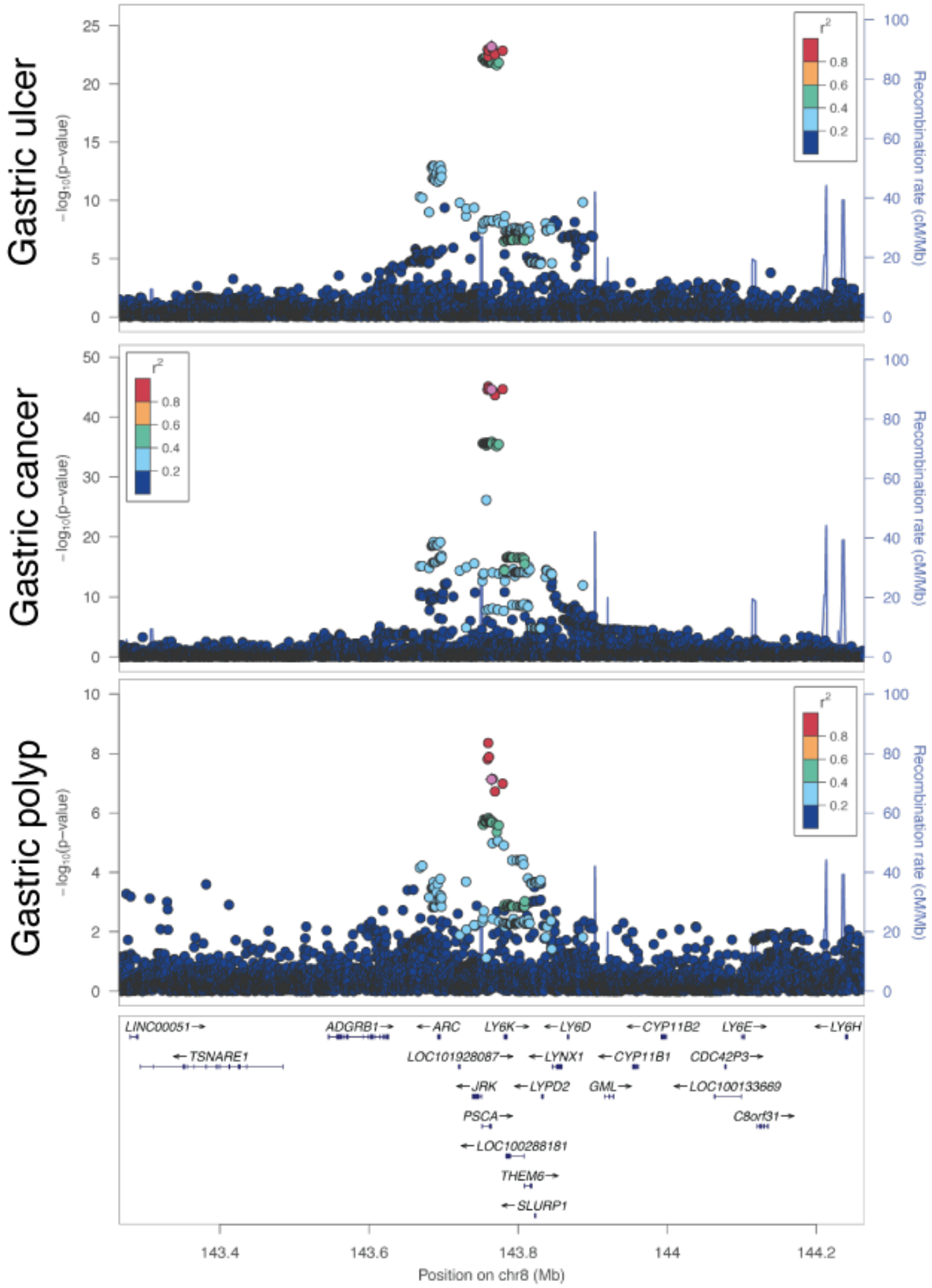


**Extended Data Figure 4 | Locus plot for gastric diseases in BBJ.**

Regional association plots at the *PSCA* locus in gastric ulcer, gastric cancer, and gastric polyp are shown. Rs2976397, which was a lead variant in gastric ulcer, is colored in pink, and colors of other dots indicate linkage disequilibrium measure $r^2$ with the lead variant.

**Extended Data Figure 5.**

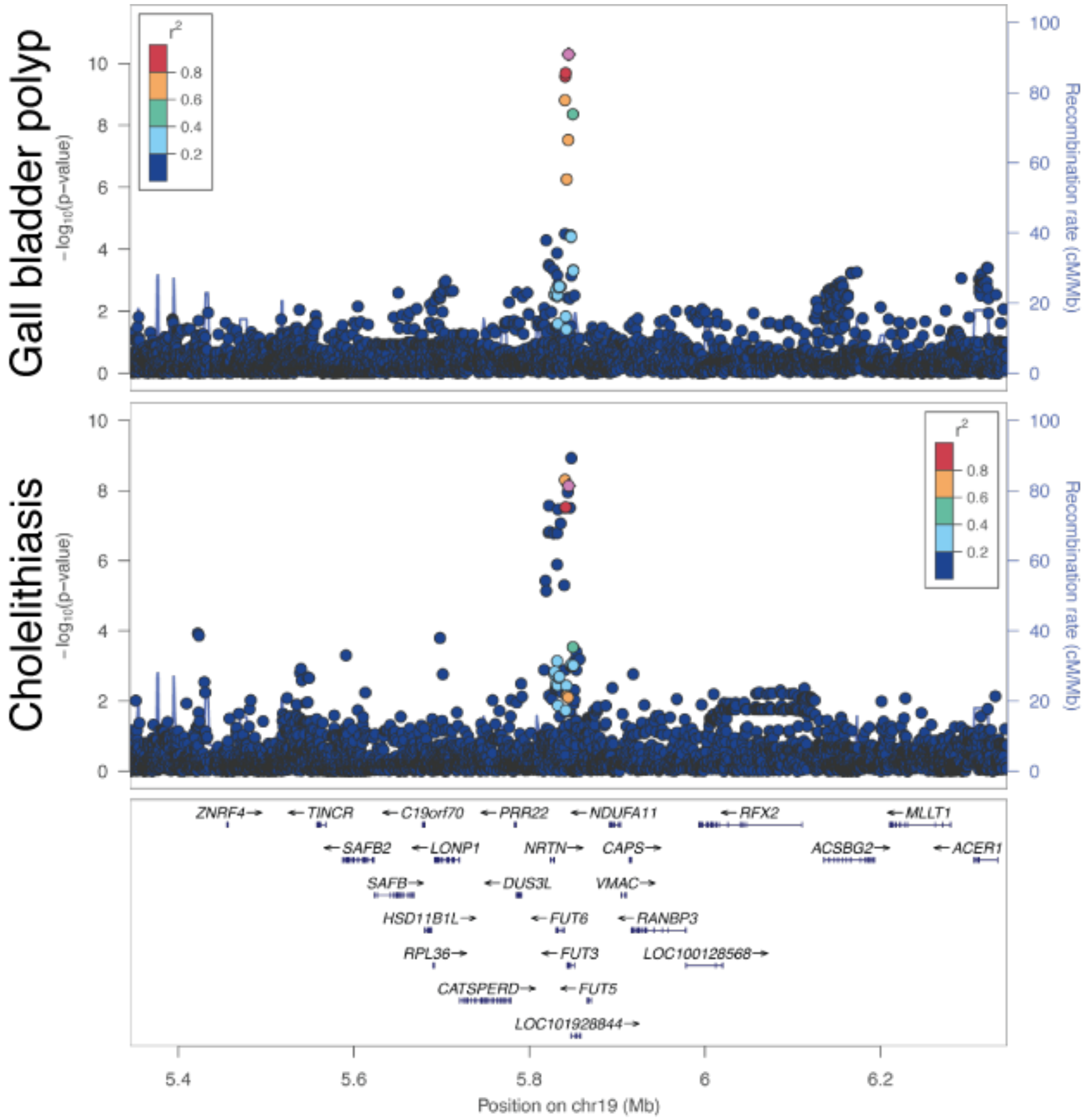


**Extended Data Figure 5 | Locus plot for gall bladder diseases in BBJ.**

Regional association plots at the *FUT3* locus in gall bladder polyp and cholelithiasis are shown. Rs28362459, which was a lead variant in gall bladder polyp, is colored in pink, and colors of other dots indicate linkage disequilibrium measure $r^2$ with the lead variant.

**Extended Data Figure 6.**

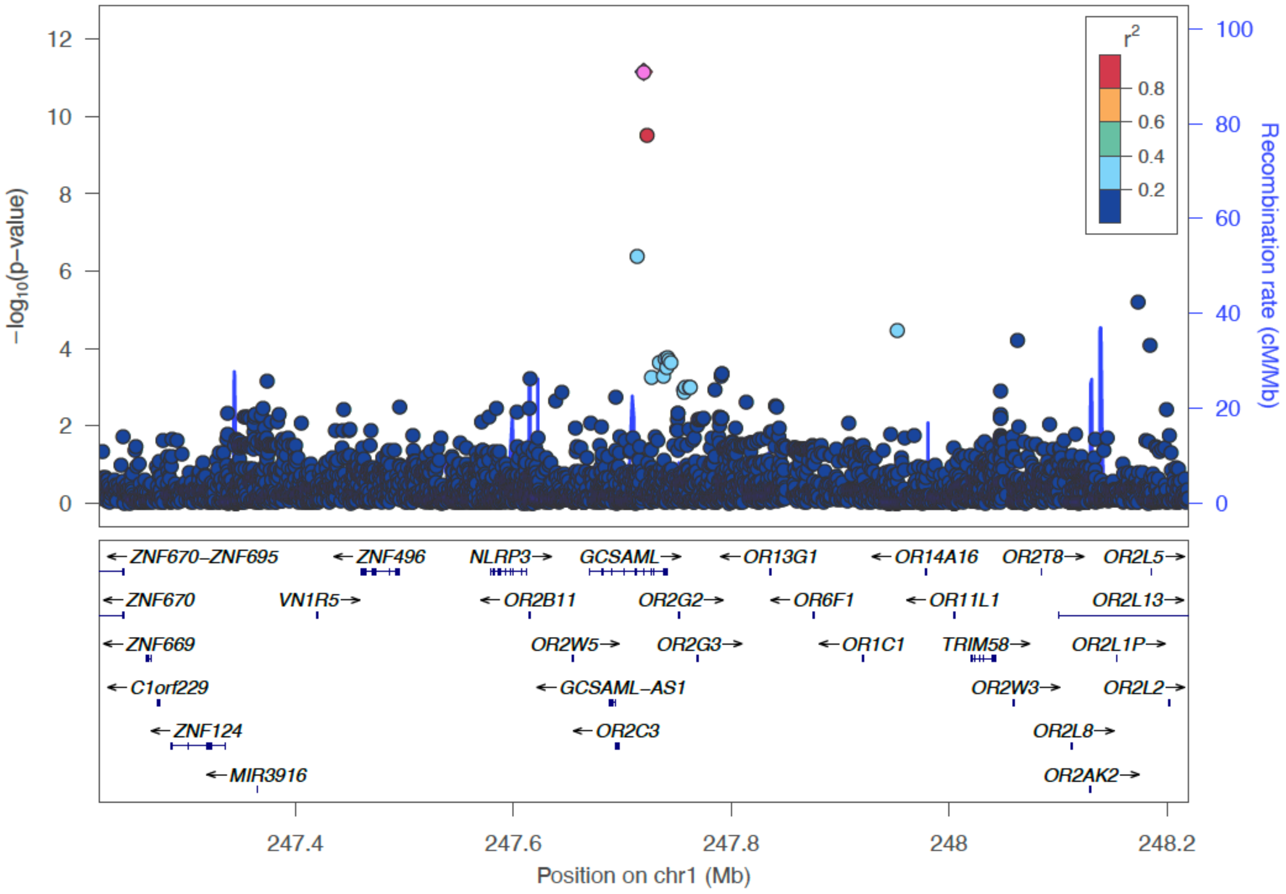


**Extended Data Figure 6 | Locus plot for urticaria in BBJ.**

Regional association plots are shown. The lead variant (rs56043070) is colored in pink, and colors of other dots indicate linkage disequilibrium measure $r^2$ with the lead variant.

**Extended Data Figure 7.**

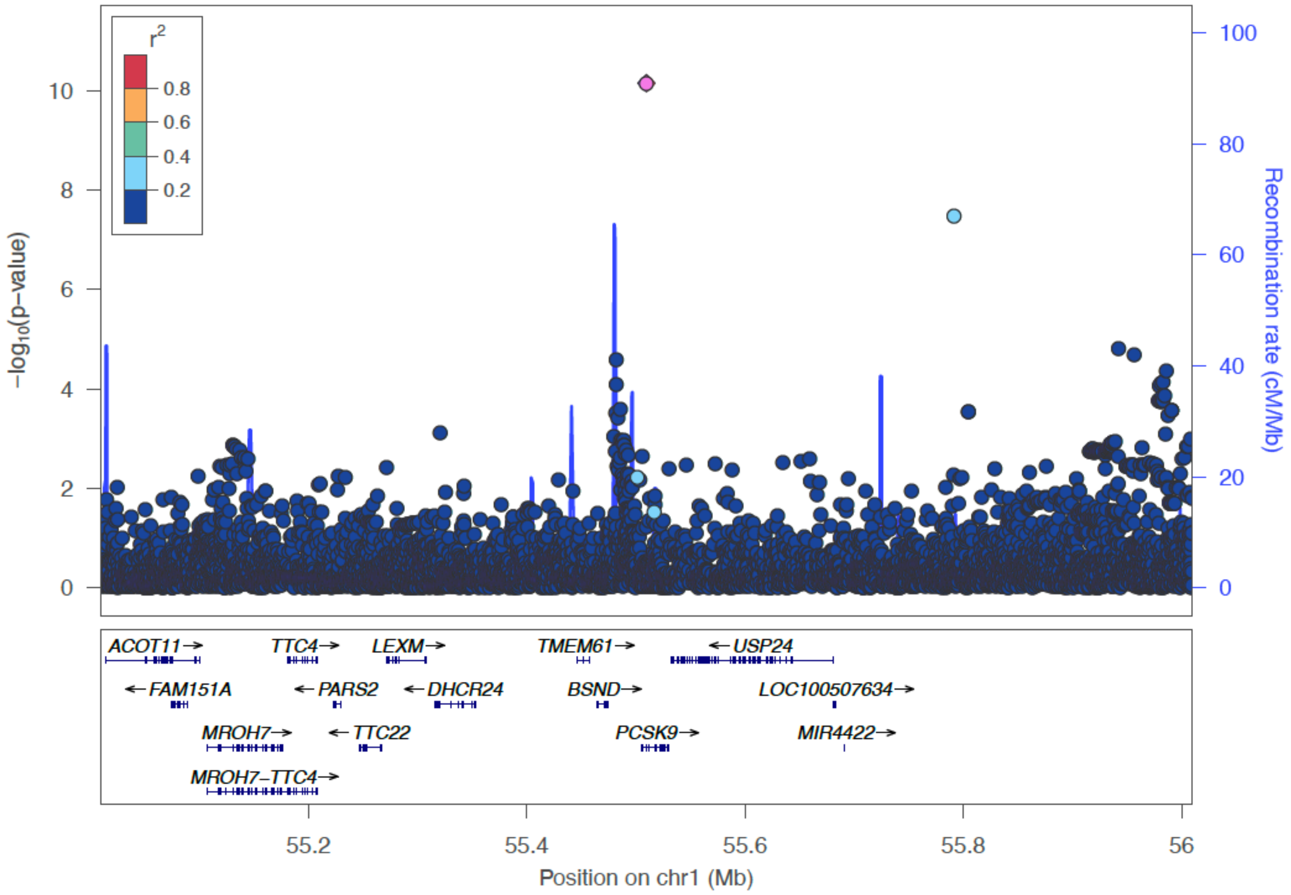


**Extended Data Figure 7 | Locus plot for salicylic acids prescription in BBJ.**

Regional association plots are shown. The lead variant (rs151193009) is colored in pink, and colors of other dots indicate linkage disequilibrium measure $r^2$ with the lead variant.

**Extended Data Figure 8.**

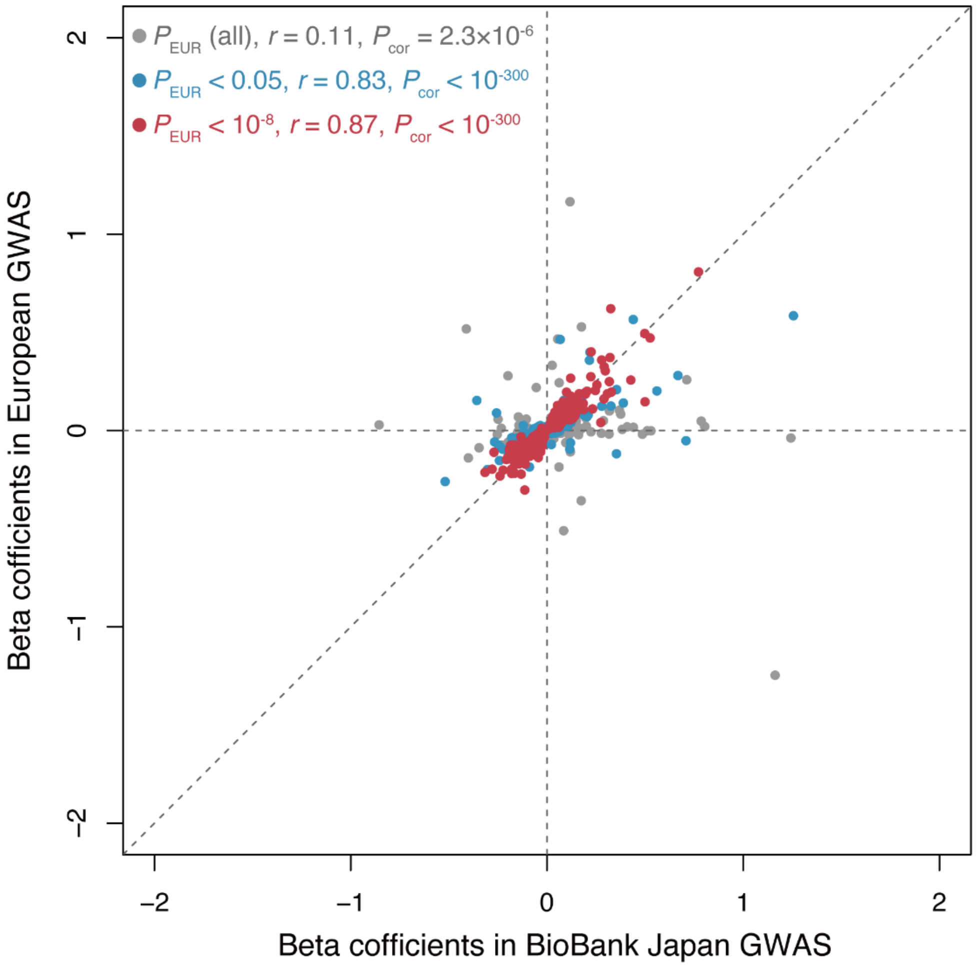


**Extended Data Figure 8 | The effect size correlation between BBJ GWAS and European GWAS.** The marginal effect sizes of genome-wide significant variants across traits (diseases and biomarkers) in BBJ are compared with those in European GWAS. Each plot represents a variant, and is colored based on the significance in European GWAS as shown in the left top legend. Pearson's correlation *r* and *P* value between BBJ GWAS and European GWAS are also shown in the legend.

**Extended Data Figure 9.**

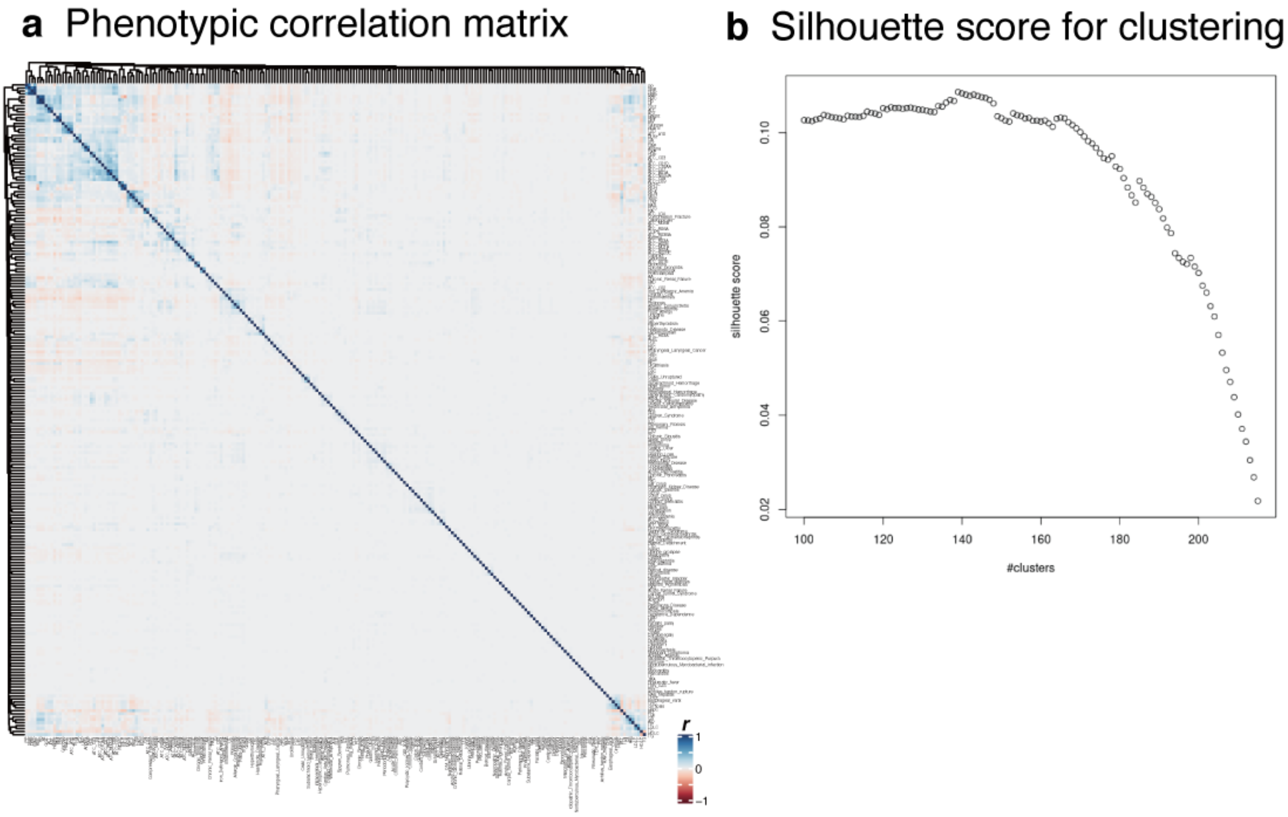


**Extended Data Figure 9 | Phenotypic correlation across 220 phenotypes in BBJ**

**a**. Heatmap of pair-wise phenotypic correlation matrix. The color of the cells indicates the value of correlation *r* as shown in a color scale at the bottom. The traits (rows and columns) were hierarchically clustered by hclust library in R. **b**. Silhouette score for clustering of closely related phenotypes with different number of clusters (**Supplementary Notes**).

**Extended Data Figure 10.**

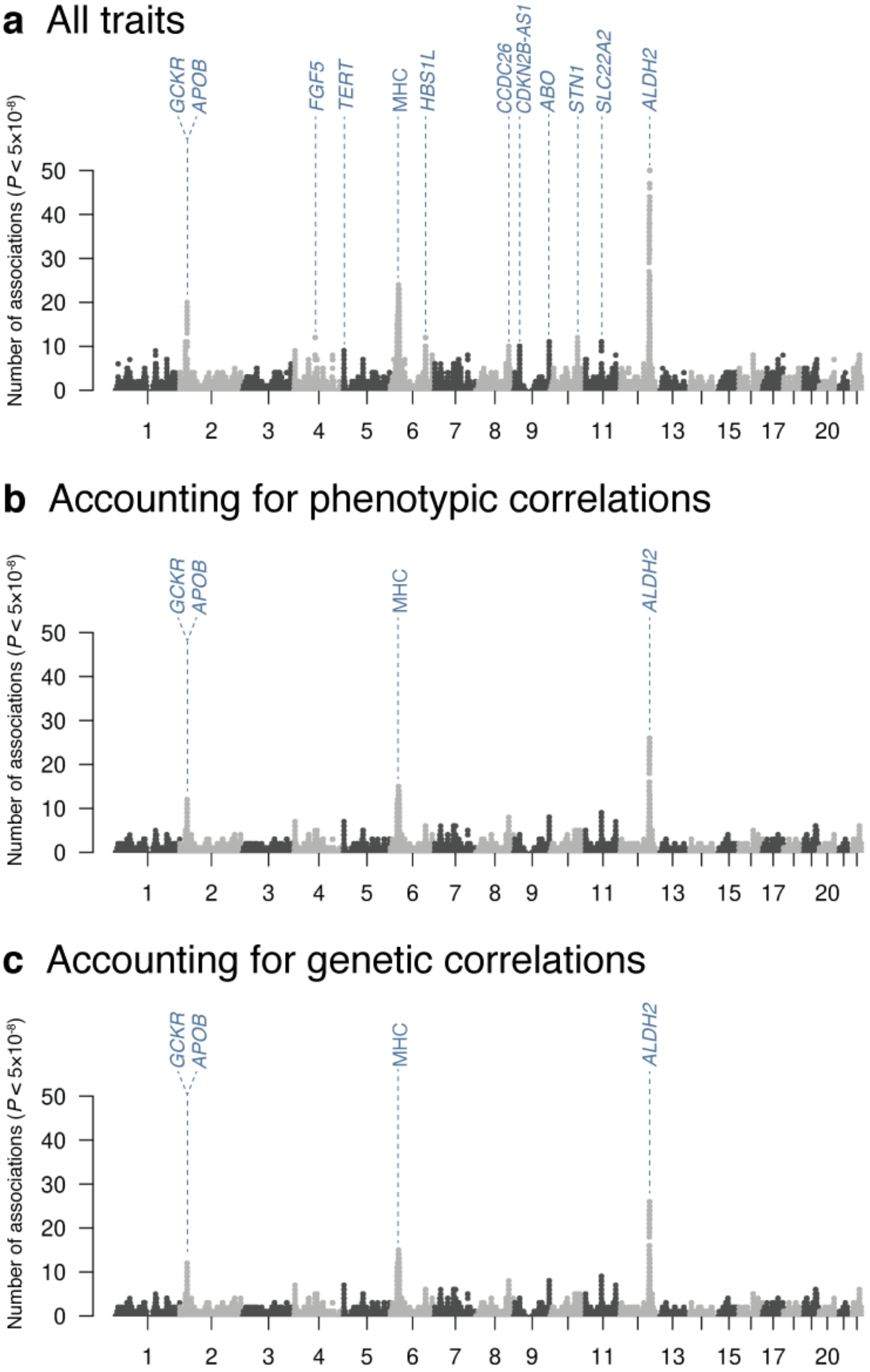


**Extended Data Figure 10 | The degree of pleiotropy in BBJ after accounting for phenotypic or genetic correlations**

The Manhattan-like plots show the number of significant associations ($P < 5.0\times10^{-8}$) at each tested genetic variant in Japanese. **a**. For all traits ($n_{\mathrm{trait}}$ = 220; as shown in Figure 2a). **b**. After accounting for phenotypic correlations. **c**. After accounting for genetic correlations.

**Extended Data Figure 11.**

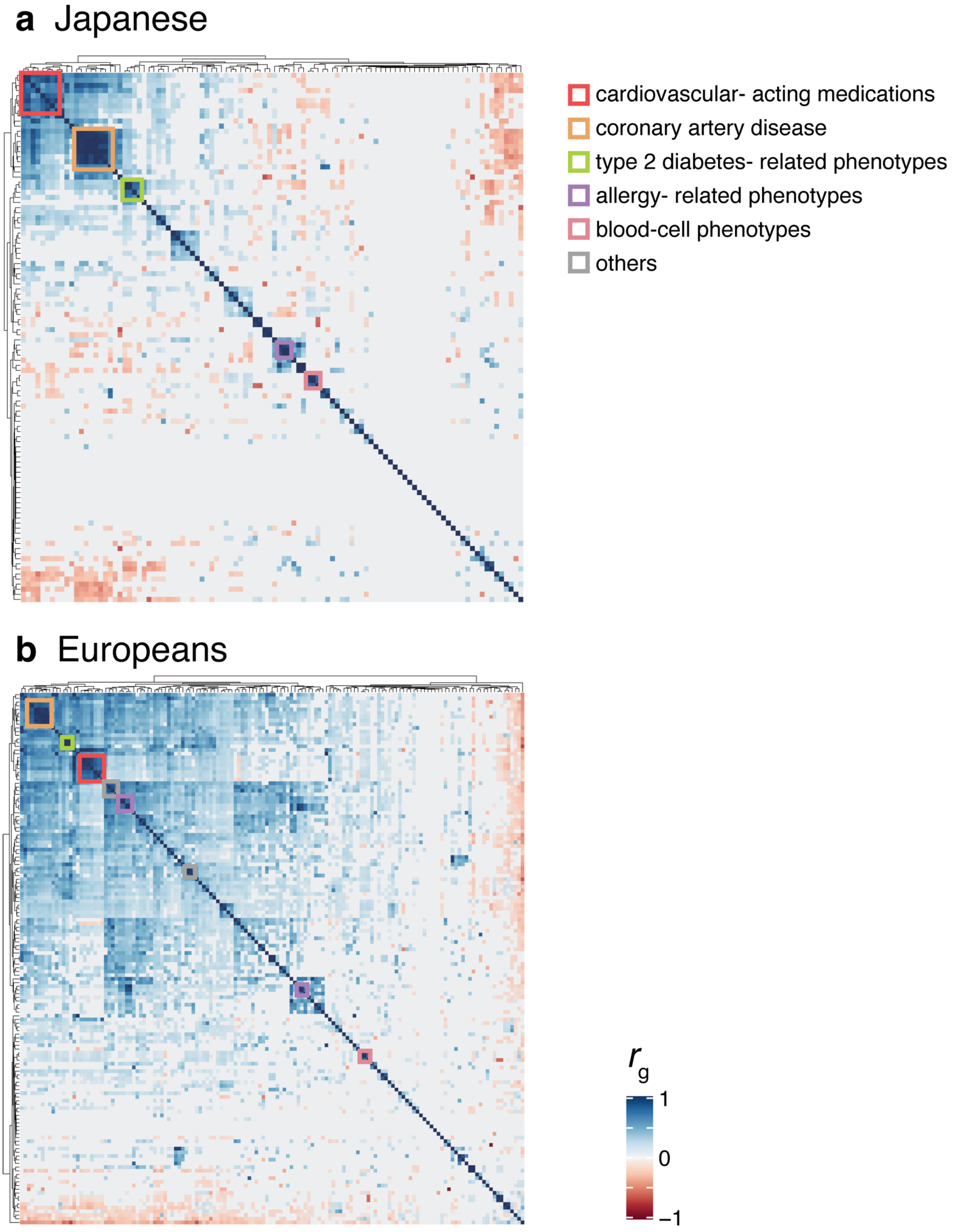


**Extended Data Figure 11 | Genetic correlation matrices across populations.**

The matrices describe pairwise genetic correlation $r_g$s in Japanese GWAS (**a**; *n* = 5,565) and in European GWAS (**b**; *n* = 10,878), which were estimated by bivariate LD score regression. A color of the cells indicates the value of $r_g$ as shown in a color scale at the bottom. The traits (rows and columns) were hierarchically clustered by hclust library in R, and trait domains are displayed as colored boxes (see **Methods**).

**Extended Data Figure 12.**

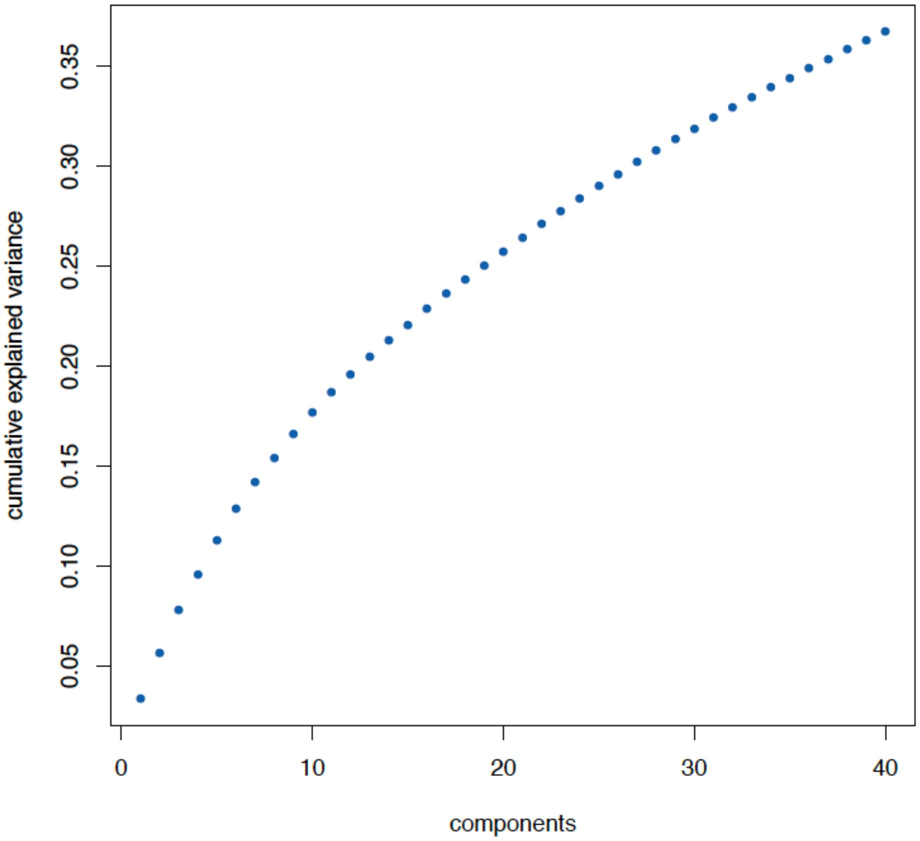


**Extended Data Figure 12 | A scree plot from the decomposition of summary statistics.**
A scree plot summarizes cumulative explained variance by the 40 components from TSVD. We calculated variance explained by each component from the ".explained_variance_ratio_" from TruncatedSVD module in python package.

**Extended Data Figure 13.**

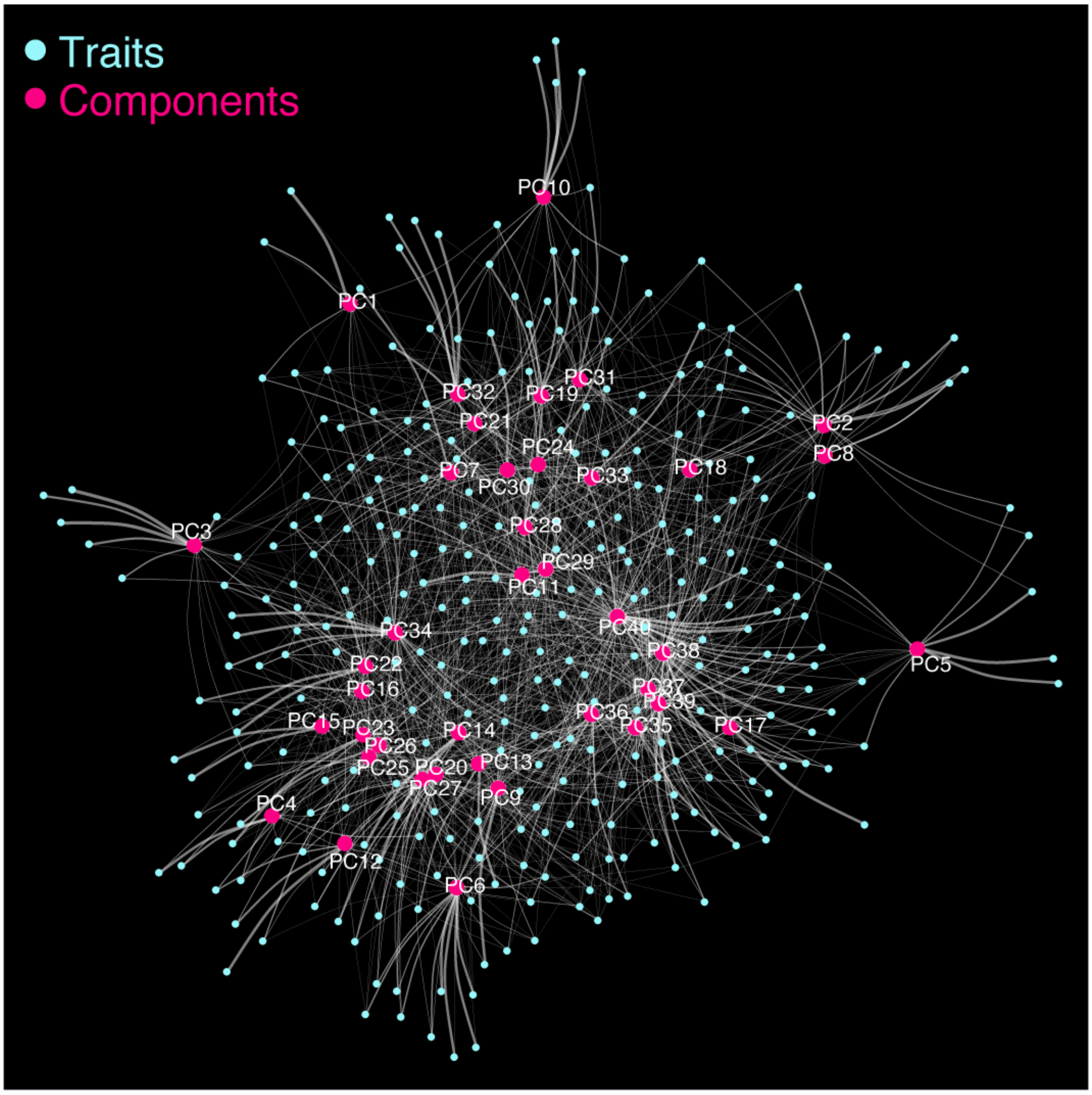


**Extended Data Figure 13 | Network representation of the TSVD analysis.**

Two-dimensional illustration of interconnection among 159 diseases and 40 latent components. Plots in blue indicate each trait's statistics, and plots in pink indicate the latent components derived by TSVD. White lines represent the contribution of each phenotype in each component. The width of the lines indicates the strength of the contribution based on the squared cosine score.

**Extended Data Figure 14.**

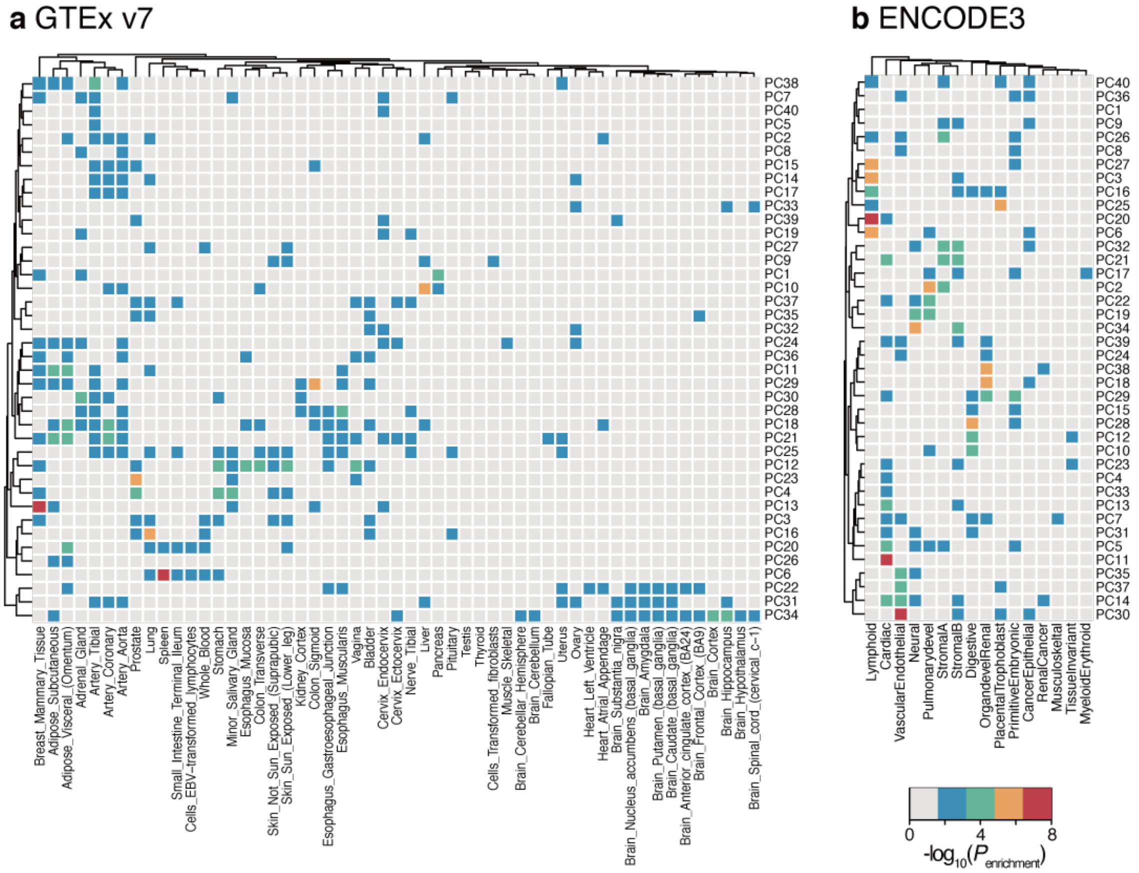


**Extended Data Figure 14 | Enrichment analyses of genes explaining each component with tissue specificity.**

A heatmap representation of the enrichment analyses of genes explaining each component with tissue-specific genes defined by GTEx expression profile (**a**) and regulatory vocabulary from ENCODE3 data (**b**). Each cell is colored based on $P_{enrichment}$ from Fisher's exact tests to assess the enrichment of the genes comprising each component within each tissue-specific gene set as shown in a color scale at the bottom right.

**Extended Data Figure 15.**

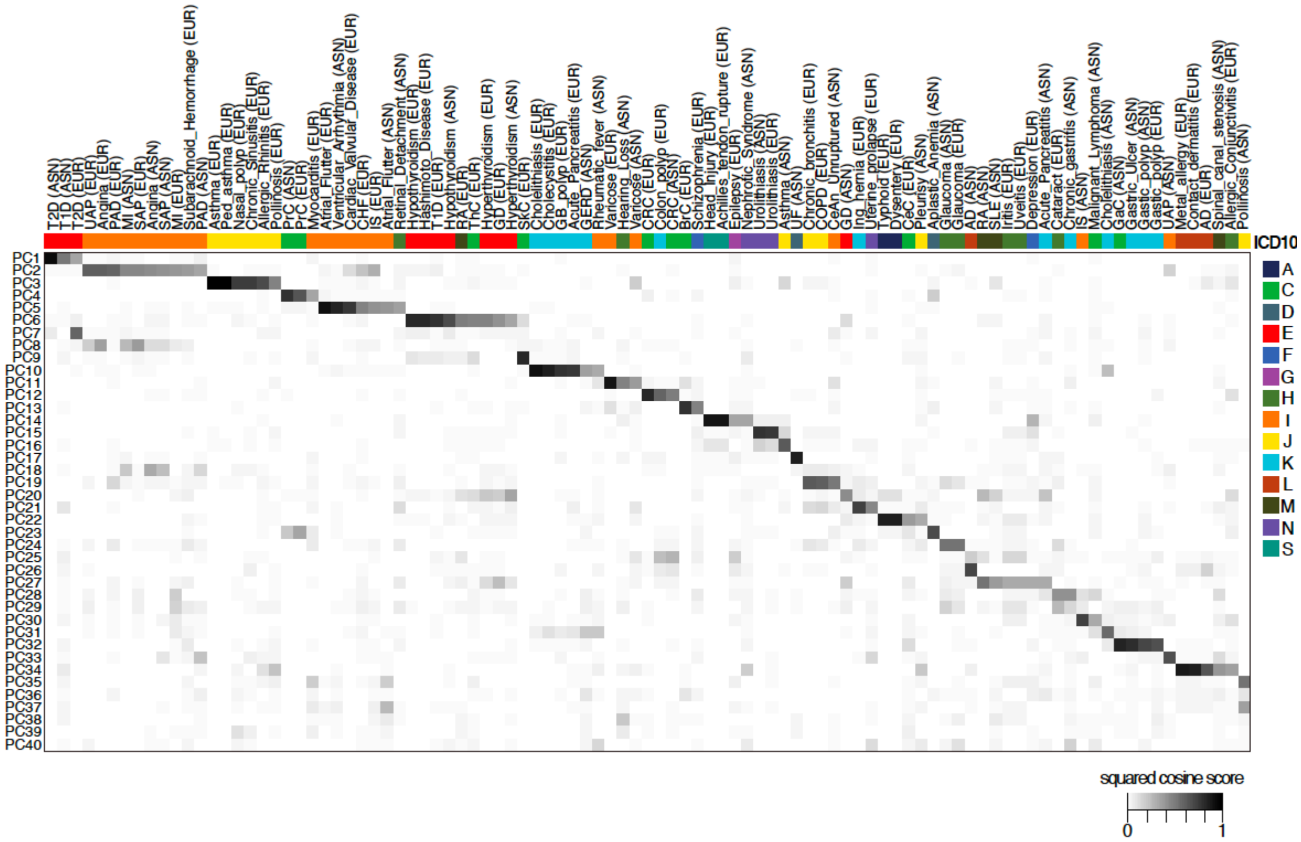


**Extended Data Figure 15 | Heatmap representation of squared cosine scores of diseases to components.**

The components (rows) are shown from 1 (top) to 40 (bottom), and the diseases (columns) are sorted based on the contribution of each component to the disease based on the squared cosine score (from component 1 to 40). Each cell is colored based on the squared cosine score of a given trait to a given component, as shown in a color scale at the bottom right.

**Extended Data Figure 16.**

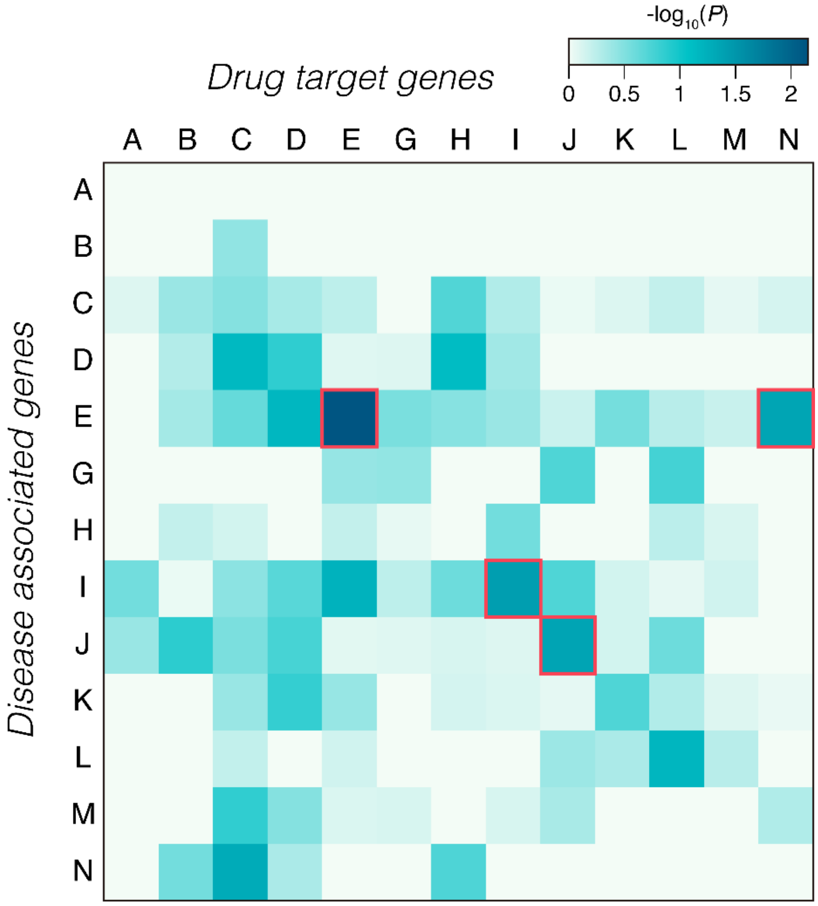


**Extended Data Figure 16 | Drug target enrichment analysis.**

The enrichment *P* value from Fisher's exact test of disease-associated genes against target genes of approved drugs for the disease is illustrated as a heatmap. The color of each cell indicates the $-\log_{10}(P)$ value of the enrichment of associated genes with each disease category to target genes of approved medication to a given disease category as shown in the color scale (top right). The disease category is ordered as an alphabetical code of the ICD10 classifications. The enrichment with nominal significance ($P < 0.05$) is highlighted as a box with a red border.

**Extended Data Figure 17.**

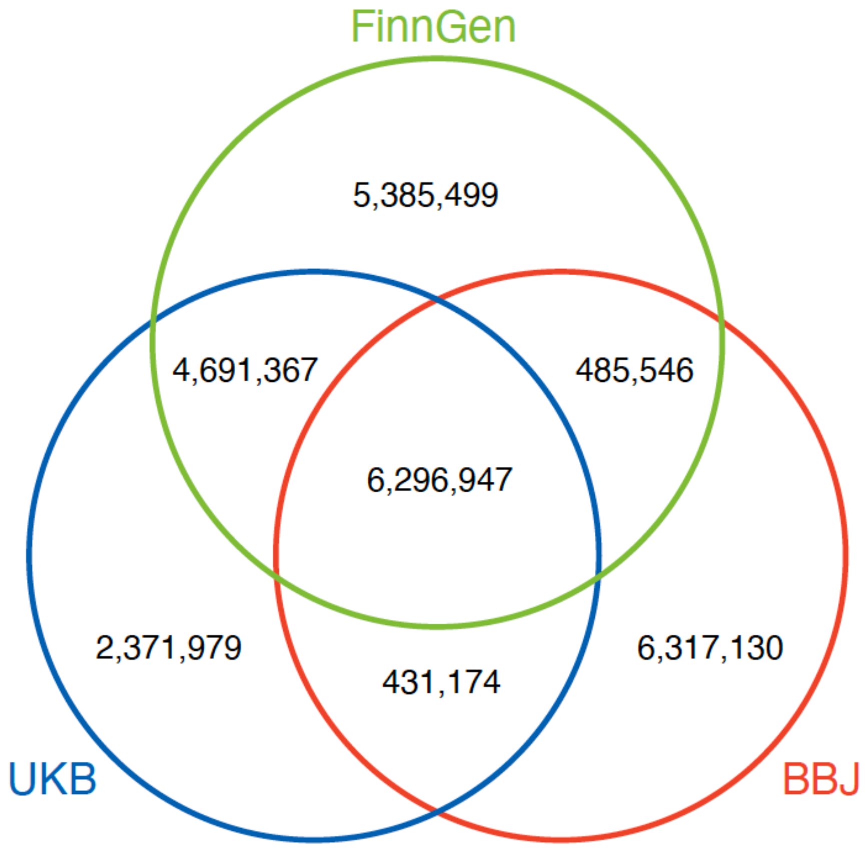


**Extended Data Figure 17 | Genetic variants analyzed in the three cohorts.**

The Venn diagram showing the number of genetic variants analyzed in this study in each of the three cohorts (BBJ, UKB, and FinnGen) and overlapping variants across the cohorts.

**Supplementary Notes**

***1. Cohort descriptions***

*The Biobank Japan*

The BioBank Japan (BBJ) Project is a nation-wide hospital-based prospective cohort launched in 2003. The BBJ collaboratively collected serum as well as clinical data of approximately 200,000 participants from 66 hospitals affiliated with 12 medical institutes. Participants were registered to the cohort from June 2003 to March 2008, and their clinical information was collected annually via interviews and medical record reviews until 2013. We collected DNA from all participants at baseline and collected annual serum samples until 2013.

*The UK Biobank*

The UK Biobank project is a population-based prospective cohort that recruited approximately 500,000 people aged between 40–69 years from 2006 to 2010 from across the United Kingdom. Deep phenotype data, such as electronic medical records, lifestyle indicators and bioassays (e.g., biochemistry, proteomic and metabonomic analyses), and genotype data were available for most of the participants. The detailed characteristics of the cohort were extensively described elsewhere[1].

*FinnGen*

FinnGen is a public–private partnership project. Six regional and three country-wide Finnish biobanks participate in FinnGen. Additionally, data from previously established population and disease-based cohorts are utilized. Participants' health outcomes are followed up by linking to the national health registries (1969–2016), which collect information from birth to death. A list of FinnGen contributors is presented below.

*The Tohoku Medical Megabank (TMM)*

The Tohoku Medical Megabank (TMM) Project was launched for the purpose of reconstruction from the Great East Japan Earthquake on 11 March 2011 and establishment of personalized healthcare and medicine[2]. In the project, an integrated biobank has been established based on two prospective genome cohort studies ("the TMM Community-Based Cohort Study" and "Birth and Three-Generation Cohort Study"). Although the cohort studies recruited residents in Miyagi or Iwate Prefecture, only the participants living in Miyagi Prefecture, whose measurements on metabolites were available, were used in the study.

***2. Evaluation of regional pleiotropy after accounting for phenotypic and genetic correlations***

When traits are phenotypically or genetically dependent, the number of closely related traits might affect the level of pleiotropy by overestimating pleiotropy at the loci harboring genetic associations with a large number of closely related traits. To evaluate the potential effect of phenotypically or genetically correlated traits on the quantified degree of pleiotropy, we sought to assess the degree of pleiotropy after accounting for phenotypic and genetic correlations. First, to account for the phenotypic correlations, we defined phenotypically correlated trait domains as in Watanabe et al[3]. Briefly, we extracted 220 BBJ trait values per individual and computed the phenotypic correlation ($r_p$) as the Pearson's correlation (**Extended Data Figure 9a**). We replaced non-significant correlation ($r_p$) between pairs of traits after Bonferroni correction of multiple testing ($P > 0.05/220$) with zero. We next clustered traits based on the phenotypic correlations using hierarchical clustering ("hclust" in R) by optimizing the number of clusters $k$ while maximizing silhouette score. In our case, at $k = 139$ the silhouette score was maximized (**Extended Data Figure 9b**) and we defined closely-related trait domains using this clustering result. Finally, we re-quantified the degree of pleiotropy by aggregating and counting the number of genome-wide significant associations across these trait domains (i.e., the association within a trait domain with multiple traits will be counted only once; **Extended Data Figure 10b**). For the definition of genetically closely related traits, we adopted the trait domains based on the genetic correlation matrix of $r_g$ for consistency (see **Methods**). Then, we re-quantified the degree of pleiotropy by aggregating and counting the number of genome-wide significant associations across the genetically-defined trait domains (**Extended Data Figure 10c**).

#### *3. Statistical fine-mapping of the top pleiotropic loci in Biobank Japan (ALDH2 and GCKR)*

We retrieved statistical fine-mapping results of the top pleiotropic loci (*ALDH2* and *GCKR*) from 67 overlapping traits studied in our ongoing study (Kanai et al., unpublished data). We used the same fine-mapping pipeline as previously described (Atkinson et al.[4] and https://www.finucanelab.org/data). Briefly, we used FINEMAP version 1.3.1[5] and susieR version 0.8.1.0521[6] for fine-mapping. As input, we used the GWAS summary statistics and in-sample dosage linkage disequilibrium (LD) matrices computed by LDstore version 2.0b. We defined regions based on 3 Mb window around lead variants and merged them if they overlapped. The maximum number of causal variants in a region was specified as ten.

We found that out of 36 and 18 traits in which rs671 (*ALDH2*) and rs1260326 (*GCKR*) are genome-wide significant ($P < 5.0 \times 10^{-8}$), 36 and 16 traits have the variant in 95% credible set, respectively. For the remaining two *GCKR* traits, we confirmed there are no additional independent signals in the locus. Altogether, these results suggest that our current approach is sufficient for the top two pleiotropic loci we identified. Further fine-mapping study should be warranted to elucidate a global landscape of pleiotropic variants in the future.

#### *4. Contributors of FinnGen*

**Steering Committee**

| | |
|---|---|
| Aarno Palotie | Institute for Molecular Medicine Finland, HiLIFE, University of Helsinki, Finland |
| Mark Daly | Institute for Molecular Medicine Finland, HiLIFE, University of Helsinki, Finland |

**Pharmaceutical companies**

| | |
|---|---|
| Howard Jacob | Abbvie, Chicago, IL, United States |
| Athena Matakidou | Astra Zeneca, Cambridge, United Kingdom |
| Heiko Runz | Biogen, Cambridge, MA, United States |
| Sally John | Biogen, Cambridge, MA, United States |
| Robert Plenge | Celgene, Summit, NJ, United States |
| Mark McCarthy | Genentech, San Francisco, CA, United States |

Julie Hunkapiller Genentech, San Francisco, CA, United States
Meg Ehm GlaxoSmithKline, Brentford, United Kingdom
Dawn Waterworth GlaxoSmithKline, Brentford, United Kingdom
Caroline Fox Merck, Kenilworth, NJ, United States
Anders Malarstig Pfizer, New York, NY, United States
Kathy Klinger Sanofi, Paris, France
Kathy Call Sanofi, Paris, France

**University of Helsinki & Biobanks**
Tomi Mäkelä HiLIFE, University of Helsinki, Finland, Finland
Jaakko Kaprio Institute for Molecular Medicine Finland, HiLIFE, Helsinki, Finland, Finland
Petri Virolainen Auria Biobank / Univ. of Turku / Hospital District of Southwest Finland, Turku, Finland
Kari Pulkki Auria Biobank / Univ. of Turku / Hospital District of Southwest Finland, Turku, Finland
Terhi Kilpi THL Biobank / The National Institute of Health and Welfare Helsinki, Finland
Markus Perola THL Biobank / The National Institute of Health and Welfare Helsinki, Finland
Jukka Partanen Finnish Red Cross Blood Service / Finnish Hematology Registry and Clinical Biobank, Helsinki, Finland
Anne Pitkäranta Hospital District of Helsinki and Uusimaa, Helsinki, Finland
Riitta Kaarteenaho Northern Finland Biobank Borealis / University of Oulu / Northern Ostrobothnia Hospital District, Oulu, Finland
Seppo Vainio Northern Finland Biobank Borealis / University of Oulu / Northern Ostrobothnia Hospital District, Oulu, Finland
Kimmo Savinainen Finnish Clinical Biobank Tampere / University of Tampere / Pirkanmaa Hospital District, Tampere, Finland
Veli-Matti Kosma Biobank of Eastern Finland / University of Eastern Finland / Northern Savo Hospital District, Kuopio, Finland
Urho Kujala Central Finland Biobank / University of Jyväskylä / Central Finland Health Care District, Jyväskylä, Finland

**Other Experts/ Non-Voting Members**
Outi Tuovila Business Finland, Helsinki, Finland
Minna Hendolin Business Finland, Helsinki, Finland
Raimo Pakkanen Business Finland, Helsinki, Finland

**Scientific Committee**
**Pharmaceutical companies**
Jeff Waring Abbvie, Chicago, IL, United States
Bridget Riley-Gillis Abbvie, Chicago, IL, United States
Athena Matakidou Astra Zeneca, Cambridge, United Kingdom
Heiko Runz Biogen, Cambridge, MA, United States
Jimmy Liu Biogen, Cambridge, MA, United States
Shameek Biswas Celgene, Summit, NJ, United States
Julie Hunkapiller Genentech, San Francisco, CA, United States
Dawn Waterworth GlaxoSmithKline, Brentford, United Kingdom
Meg Ehm GlaxoSmithKline, Brentford, United Kingdom
Dorothee Diogo Merck, Kenilworth, NJ, United States
Caroline Fox Merck, Kenilworth, NJ, United States
Anders Malarstig Pfizer, New York, NY, United States
Catherine Marshall Pfizer, New York, NY, United States
Xinli Hu Pfizer, New York, NY, United States
Kathy Call Sanofi, Paris, France
Kathy Klinger Sanofi, Paris, France
Matthias Gossel Sanofi, Paris, France

**University of Helsinki & Biobanks**
Samuli Ripatti Institute for Molecular Medicine Finland, HiLIFE, Helsinki, Finland
Johanna Schleutker Auria Biobank / Univ. of Turku / Hospital District of Southwest Finland, Turku, Finland

| | |
|---|---|
| Markus Perola | THL Biobank / The National Institute of Health and Welfare Helsinki, Finland |
| Mikko Arvas | Finnish Red Cross Blood Service / Finnish Hematology Registry and Clinical Biobank, Helsinki, Finland |
| Olli Carpen | Hospital District of Helsinki and Uusimaa, Helsinki, Finland |
| Reetta Hinttala | Northern Finland Biobank Borealis / University of Oulu / Northern Ostrobothnia Hospital District, Oulu, Finland |
| Johannes Kettunen | Northern Finland Biobank Borealis / University of Oulu / Northern Ostrobothnia Hospital District, Oulu, Finland |
| Reijo Laaksonen | Finnish Clinical Biobank Tampere / University of Tampere / Pirkanmaa Hospital District, Tampere, Finland |
| Arto Mannermaa | Biobank of Eastern Finland / University of Eastern Finland / Northern Savo Hospital District, Kuopio, Finland |
| Juha Paloneva | Central Finland Biobank / University of Jyväskylä / Central Finland Health Care District, Jyväskylä, Finland |
| Urho Kujala | Central Finland Biobank / University of Jyväskylä / Central Finland Health Care District, Jyväskylä, Finland |

**Other Experts/ Non-Voting Members**

| | |
|---|---|
| Outi Tuovila | Business Finland, Helsinki, Finland |
| Minna Hendolin | Business Finland, Helsinki, Finland |
| Raimo Pakkanen | Business Finland, Helsinki, Finland |

**Clinical Groups**

**Neurology Group**

| | |
|---|---|
| Hilkka Soininen | Northern Savo Hospital District, Kuopio, Finland |
| Valtteri Julkunen | Northern Savo Hospital District, Kuopio, Finland |
| Anne Remes | Northern Ostrobothnia Hospital District, Oulu, Finland |
| Reetta Kälviäinen | Northern Savo Hospital District, Kuopio, Finland |
| Mikko Hiltunen | Northern Savo Hospital District, Kuopio, Finland |
| Jukka Peltola | Pirkanmaa Hospital District, Tampere, Finland |
| Pentti Tienari | Hospital District of Helsinki and Uusimaa, Helsinki, Finland |
| Juha Rinne | Hospital District of Southwest Finland, Turku, Finland |
| Adam Ziemann | Abbvie, Chicago, IL, United States |
| Jeffrey Waring | Abbvie, Chicago, IL, United States |
| Sahar Esmaeeli | Abbvie, Chicago, IL, United States |
| Nizar Smaoui | Abbvie, Chicago, IL, United States |
| Anne Lehtonen | Abbvie, Chicago, IL, United States |
| Susan Eaton | Biogen, Cambridge, MA, United States |
| Heiko Runz | Biogen, Cambridge, MA, United States |
| Sanni Lahdenperä | Biogen, Cambridge, MA, United States |
| Shameek Biswas | Celgene, Summit, NJ, United States |
| John Michon | Genentech, San Francisco, CA, United States |
| Geoff Kerchner | Genentech, San Francisco, CA, United States |
| Julie Hunkapiller | Genentech, San Francisco, CA, United States |
| Natalie Bowers | Genentech, San Francisco, CA, United States |
| Edmond Teng | Genentech, San Francisco, CA, United States |
| John Eicher | Merck, Kenilworth, NJ, United States |
| Vinay Mehta | Merck, Kenilworth, NJ, United States |
| Padhraig Gormley | Merck, Kenilworth, NJ, United States |
| Kari Linden | Pfizer, New York, NY, United States |
| Christopher Whelan | Pfizer, New York, NY, United States |
| Fanli Xu | GlaxoSmithKline, Brentford, United Kingdom |
| David Pulford | GlaxoSmithKline, Brentford, United Kingdom |

**Gastroenterology Group**

| | |
|---|---|
| Martti Färkkilä | Hospital District of Helsinki and Uusimaa, Helsinki, Finland |
| | Sampsa Pikkarainen Hospital District of Helsinki and Uusimaa, Helsinki, Finland |
| Airi Jussila | Pirkanmaa Hospital District, Tampere, Finland |

| | |
|---|---|
| Timo Blomster | Northern Ostrobothnia Hospital District, Oulu, Finland |
| Mikko Kiviniemi | Northern Savo Hospital District, Kuopio, Finland |
| | Markku Voutilainen Hospital District of Southwest Finland, Turku, Finland |
| Bob Georgantas | Abbvie, Chicago, IL, United States |
| Graham Heap | Abbvie, Chicago, IL, United States |
| Jeffrey Waring | Abbvie, Chicago, IL, United States |
| Nizar Smaoui | Abbvie, Chicago, IL, United States |
| Fedik Rahimov | Abbvie, Chicago, IL, United States |
| Anne Lehtonen | Abbvie, Chicago, IL, United States |
| Keith Usiskin | Celgene, Summit, NJ, United States |
| Joseph Maranville | Celgene, Summit, NJ, United States |
| Tim Lu | Genentech, San Francisco, CA, United States |
| Natalie Bowers | Genentech, San Francisco, CA, United States |
| Danny Oh | Genentech, San Francisco, CA, United States |
| John Michon | Genentech, San Francisco, CA, United States |
| Vinay Mehta | Merck, Kenilworth, NJ, United States |
| Kirsi Kalpala | Pfizer, New York, NY, United States |
| Melissa Miller | Pfizer, New York, NY, United States |
| Xinli Hu | Pfizer, New York, NY, United States |
| Linda McCarthy | GlaxoSmithKline, Brentford, United Kingdom |

**Rheumatology Group**

| | |
|---|---|
| Kari Eklund | Hospital District of Helsinki and Uusimaa, Helsinki, Finland |
| Antti Palomäki | Hospital District of Southwest Finland, Turku, Finland |
| Pia Isomäki | Pirkanmaa Hospital District, Tampere, Finland |
| Laura Pirilä | Hospital District of Southwest Finland, Turku, Finland |
| Oili Kaipiainen-Seppänen | Northern Savo Hospital District, Kuopio, Finland |
| Johanna Huhtakangas | Northern Ostrobothnia Hospital District, Oulu, Finland |
| Bob Georgantas | Abbvie, Chicago, IL, United States |
| Jeffrey Waring | Abbvie, Chicago, IL, United States |
| Fedik Rahimov | Abbvie, Chicago, IL, United States |
| | Apinya LertratanakulAbbvie, Chicago, IL, United States |
| Nizar Smaoui | Abbvie, Chicago, IL, United States |
| Anne Lehtonen | Abbvie, Chicago, IL, United States |
| David Close | Astra Zeneca, Cambridge, United Kingdom |
| Marla Hochfeld | Celgene, Summit, NJ, United States |
| Natalie Bowers | Genentech, San Francisco, CA, United States |
| John Michon | Genentech, San Francisco, CA, United States |
| Dorothee Diogo | Merck, Kenilworth, NJ, United States |
| Vinay Mehta | Merck, Kenilworth, NJ, United States |
| Kirsi Kalpala | Pfizer, New York, NY, United States |
| Nan Bing | Pfizer, New York, NY, United States |
| Xinli Hu | Pfizer, New York, NY, United States |
| Jorge Esparza Gordillo | GlaxoSmithKline, Brentford, United Kingdom |
| Nina Mars | Institute for Molecular Medicine Finland, HiLIFE, Helsinki, Finland |

**Pulmonology Group**

| | |
|---|---|
| Tarja Laitinen | Pirkanmaa Hospital District, Tampere, Finland |
| Margit Pelkonen | Northern Savo Hospital District, Kuopio, Finland |
| Paula Kauppi | Hospital District of Helsinki and Uusimaa, Helsinki, Finland |
| Hannu Kankaanranta | Pirkanmaa Hospital District, Tampere, Finland |
| Terttu Harju | Northern Ostrobothnia Hospital District, Oulu, Finland |
| Nizar Smaoui | Abbvie, Chicago, IL, United States |
| David Close | Astra Zeneca, Cambridge, United Kingdom |
| Steven Greenberg | Celgene, Summit, NJ, United States |
| Hubert Chen | Genentech, San Francisco, CA, United States |
| Natalie Bowers | Genentech, San Francisco, CA, United States |
| John Michon | Genentech, San Francisco, CA, United States |

| | |
|---|---|
| Vinay Mehta | Merck, Kenilworth, NJ, United States |
| Jo Betts | GlaxoSmithKline, Brentford, United Kingdom |
| Soumitra Ghosh | GlaxoSmithKline, Brentford, United Kingdom |

**Cardiometabolic Diseases Group**

| | |
|---|---|
| Veikko Salomaa | The National Institute of Health and Welfare Helsinki, Finland |
| Teemu Niiranen | The National Institute of Health and Welfare Helsinki, Finland |
| Markus Juonala | Hospital District of Southwest Finland, Turku, Finland |
| Kaj Metsärinne | Hospital District of Southwest Finland, Turku, Finland |
| Mika Kähönen | Pirkanmaa Hospital District, Tampere, Finland |
| Juhani Junttila | Northern Ostrobothnia Hospital District, Oulu, Finland |
| Markku Laakso | Northern Savo Hospital District, Kuopio, Finland |
| Jussi Pihlajamäki | Northern Savo Hospital District, Kuopio, Finland |
| Juha Sinisalo | Hospital District of Helsinki and Uusimaa, Helsinki, Finland |
| Marja-Riitta Taskinen | Hospital District of Helsinki and Uusimaa, Helsinki, Finland |
| Tiinamaija Tuomi | Hospital District of Helsinki and Uusimaa, Helsinki, Finland |
| Jari Laukkanen | Central Finland Health Care District, Jyväskylä, Finland |
| Ben Challis | Astra Zeneca, Cambridge, United Kingdom |
| Andrew Peterson | Genentech, San Francisco, CA, United States |
| Julie Hunkapiller | Genentech, San Francisco, CA, United States |
| Natalie Bowers | Genentech, San Francisco, CA, United States |
| John Michon | Genentech, San Francisco, CA, United States |
| Dorothee Diogo | Merck, Kenilworth, NJ, United States |
| Audrey Chu | Merck, Kenilworth, NJ, United States |
| Vinay Mehta | Merck, Kenilworth, NJ, United States |
| Jaakko Parkkinen | Pfizer, New York, NY, United States |
| Melissa Miller | Pfizer, New York, NY, United States |
| Anthony Muslin | Sanofi, Paris, France |
| Dawn Waterworth | GlaxoSmithKline, Brentford, United Kingdom |

**Oncology Group**

| | |
|---|---|
| Heikki Joensuu | Hospital District of Helsinki and Uusimaa, Helsinki, Finland |
| Tuomo Meretoja | Hospital District of Helsinki and Uusimaa, Helsinki, Finland |
| Olli Carpen | Hospital District of Helsinki and Uusimaa, Helsinki, Finland |
| Lauri Aaltonen | Hospital District of Helsinki and Uusimaa, Helsinki, Finland |
| Annika Auranen | Pirkanmaa Hospital District , Tampere, Finland |
| Peeter Karihtala | Northern Ostrobothnia Hospital District, Oulu, Finland |
| Saila Kauppila | Northern Ostrobothnia Hospital District, Oulu, Finland |
| Päivi Auvinen | Northern Savo Hospital District, Kuopio, Finland |
| Klaus Elenius | Hospital District of Southwest Finland, Turku, Finland |
| Relja Popovic | Abbvie, Chicago, IL, United States |
| Jeffrey Waring | Abbvie, Chicago, IL, United States |
| Bridget Riley-Gillis | Abbvie, Chicago, IL, United States |
| Anne Lehtonen | Abbvie, Chicago, IL, United States |
| Athena Matakidou | Astra Zeneca, Cambridge, United Kingdom |
| Jennifer Schutzman | Genentech, San Francisco, CA, United States |
| Julie Hunkapiller | Genentech, San Francisco, CA, United States |
| Natalie Bowers | Genentech, San Francisco, CA, United States |
| John Michon | Genentech, San Francisco, CA, United States |
| Vinay Mehta | Merck, Kenilworth, NJ, United States |
| Andrey Loboda | Merck, Kenilworth, NJ, United States |
| Aparna Chhibber | Merck, Kenilworth, NJ, United States |
| Heli Lehtonen | Pfizer, New York, NY, United States |
| Stefan McDonough | Pfizer, New York, NY, United States |
| Marika Crohns | Sanofi, Paris, France |
| Diptee Kulkarni | GlaxoSmithKline, Brentford, United Kingdom |

**Opthalmology Group**

| | |
|---|---|
| Kai Kaarniranta | Northern Savo Hospital District, Kuopio, Finland |
| Joni A Turunen | Hospital District of Helsinki and Uusimaa, Helsinki, Finland |
| Terhi Ollila | Hospital District of Helsinki and Uusimaa, Helsinki, Finland |
| Sanna Seitsonen | Hospital District of Helsinki and Uusimaa, Helsinki, Finland |
| Hannu Uusitalo | Pirkanmaa Hospital District, Tampere, Finland |
| Vesa Aaltonen | Hospital District of Southwest Finland, Turku, Finland |
| | Hannele Uusitalo-Järvinen Pirkanmaa Hospital District, Tampere, Finland |
| Marja Luodonpää | Northern Ostrobothnia Hospital District, Oulu, Finland |
| Nina Hautala | Northern Ostrobothnia Hospital District, Oulu, Finland |
| Heiko Runz | Biogen, Cambridge, MA, United States |
| Erich Strauss | Genentech, San Francisco, CA, United States |
| Natalie Bowers | Genentech, San Francisco, CA, United States |
| Hao Chen | Genentech, San Francisco, CA, United States |
| John Michon | Genentech, San Francisco, CA, United States |
| Anna Podgornaia | Merck, Kenilworth, NJ, United States |
| Vinay Mehta | Merck, Kenilworth, NJ, United States |
| Dorothee Diogo | Merck, Kenilworth, NJ, United States |
| Joshua Hoffman | GlaxoSmithKline, Brentford, United Kingdom |

**Dermatology Group**

| | |
|---|---|
| Kaisa Tasanen | Northern Ostrobothnia Hospital District, Oulu, Finland |
| Laura Huilaja | Northern Ostrobothnia Hospital District, Oulu, Finland |
| Katariina Hannula-Jouppi | Hospital District of Helsinki and Uusimaa, Helsinki, Finland |
| Teea Salmi | Pirkanmaa Hospital District, Tampere, Finland |
| Sirkku Peltonen | Hospital District of Southwest Finland, Turku, Finland |
| Leena Koulu | Hospital District of Southwest Finland, Turku, Finland |
| Ilkka Harvima | Northern Savo Hospital District, Kuopio, Finland |
| Kirsi Kalpala | Pfizer, New York, NY, United States |
| Ying Wu | Pfizer, New York, NY, United States |
| David Choy | Genentech, San Francisco, CA, United States |
| John Michon | Genentech, San Francisco, CA, United States |
| Nizar Smaoui | Abbvie, Chicago, IL, United States |
| Fedik Rahimov | Abbvie, Chicago, IL, United States |
| Anne Lehtonen | Abbvie, Chicago, IL, United States |
| Dawn Waterworth | GlaxoSmithKline, Brentford, United Kingdom |

**FinnGen Teams**

**Administration Team**

| | |
|---|---|
| Anu Jalanko | Institute for Molecular Medicine Finland, HiLIFE, University of Helsinki, Finland |
| Risto Kajanne | Institute for Molecular Medicine Finland, HiLIFE, University of Helsinki, Finland |
| Ulrike Lyhs | Institute for Molecular Medicine Finland, HiLIFE, University of Helsinki, Finland |

**Communication**

| | |
|---|---|
| Mari Kaunisto | Institute for Molecular Medicine Finland, HiLIFE, University of Helsinki, Finland |

**Analysis Team**

| | |
|---|---|
| Justin Wade Davis | Abbvie, Chicago, IL, United States |
| Bridget Riley-Gillis | Abbvie, Chicago, IL, United States |
| Danjuma Quarless | Abbvie, Chicago, IL, United States |
| Slavé Petrovski | Astra Zeneca, Cambridge, United Kingdom |
| Jimmy Liu | Biogen, Cambridge, MA, United States |
| Chia-Yen Chen | Biogen, Cambridge, MA, United States |
| Paola Bronson | Biogen, Cambridge, MA, United States |
| Robert Yang | Celgene, Summit, NJ, United States |
| Joseph Maranville | Celgene, Summit, NJ, United States |
| Shameek Biswas | Celgene, Summit, NJ, United States |

| | |
|---|---|
| Diana Chang | Genentech, San Francisco, CA, United States |
| Julie Hunkapiller | Genentech, San Francisco, CA, United States |
| Tushar Bhangale | Genentech, San Francisco, CA, United States |
| Natalie Bowers | Genentech, San Francisco, CA, United States |
| Dorothee Diogo | Merck, Kenilworth, NJ, United States |
| Emily Holzinger | Merck, Kenilworth, NJ, United States |
| Padhraig Gormley | Merck, Kenilworth, NJ, United States |
| Xulong Wang | Merck, Kenilworth, NJ, United States |
| Xing Chen | Pfizer, New York, NY, United States |
| Åsa Hedman | Pfizer, New York, NY, United States |
| Kirsi Auro | GlaxoSmithKline, Brentford, United Kingdom |
| Clarence Wang | Sanofi, Paris, France |
| Ethan Xu | Sanofi, Paris, France |
| Franck Auge | Sanofi, Paris, France |
| Clement Chatelain | Sanofi, Paris, France |
| Mitja Kurki | Institute for Molecular Medicine Finland, HiLIFE, University of Helsinki, Finland / Broad Institute, Cambridge, MA, United States |
| Samuli Ripatti | Institute for Molecular Medicine Finland, HiLIFE, University of Helsinki, Finland |
| Mark Daly | Institute for Molecular Medicine Finland, HiLIFE, University of Helsinki, Finland |
| Juha Karjalainen | Institute for Molecular Medicine Finland, HiLIFE, University of Helsinki, Finland / Broad Institute, Cambridge, MA, United States |
| Aki Havulinna | Institute for Molecular Medicine Finland, HiLIFE, University of Helsinki, Finland |
| Anu Jalanko | Institute for Molecular Medicine Finland, HiLIFE, University of Helsinki, Finland |
| Kimmo Palin | University of Helsinki, Helsinki, Finland |
| Priit Palta | Institute for Molecular Medicine Finland, HiLIFE, University of Helsinki, Finland |
| Pietro Della Briotta Parolo | Institute for Molecular Medicine Finland, HiLIFE, University of Helsinki, Finland |
| Wei Zhou | Broad Institute, Cambridge, MA, United States |
| Susanna Lemmelä | Institute for Molecular Medicine Finland, HiLIFE, University of Helsinki, Finland |
| Manuel Rivas | University of Stanford, Stanford, CA, United States |
| Jarmo Harju | Institute for Molecular Medicine Finland, HiLIFE, University of Helsinki, Finland |
| Aarno Palotie | Institute for Molecular Medicine Finland, HiLIFE, University of Helsinki, Finland |
| Arto Lehisto | Institute for Molecular Medicine Finland, HiLIFE, University of Helsinki, Finland |
| Andrea Ganna | Institute for Molecular Medicine Finland, HiLIFE, University of Helsinki, Finland |
| Vincent Llorens | Institute for Molecular Medicine Finland, HiLIFE, University of Helsinki, Finland |
| Antti Karlsson | Auria Biobank / Univ. of Turku / Hospital District of Southwest Finland, Turku, Finland |
| Kati Kristiansson | THL Biobank / The National Institute of Health and Welfare Helsinki, Finland |
| Mikko Arvas | Finnish Red Cross Blood Service / Finnish Hematology Registry and Clinical Biobank, Helsinki, Finland |
| Kati Hyvärinen | Finnish Red Cross Blood Service / Finnish Hematology Registry and Clinical Biobank, Helsinki, Finland |
| Jarmo Ritari | Finnish Red Cross Blood Service / Finnish Hematology Registry and Clinical Biobank, Helsinki, Finland |
| Tiina Wahlfors | Finnish Red Cross Blood Service / Finnish Hematology Registry and Clinical Biobank, Helsinki, Finland |
| Miika Koskinen | Hospital District of Helsinki and Uusimaa, Helsinki, Finland BB/HUS/Univ Hosp Districts |
| Olli Carpen | Hospital District of Helsinki and Uusimaa, Helsinki, Finland BB/HUS/Univ Hosp Districts |
| Johannes Kettunen | Northern Finland Biobank Borealis / University of Oulu / Northern Ostrobothnia Hospital District, Oulu, Finland |
| Katri Pylkäs | Northern Finland Biobank Borealis / University of Oulu / Northern Ostrobothnia Hospital District, Oulu, Finland |
| Marita Kalaoja | Northern Finland Biobank Borealis / University of Oulu / Northern Ostrobothnia Hospital District, Oulu, Finland |
| Minna Karjalainen | Northern Finland Biobank Borealis / University of Oulu / Northern Ostrobothnia Hospital District, Oulu, Finland |
| Tuomo Mantere | Northern Finland Biobank Borealis / University of Oulu / Northern Ostrobothnia Hospital District, Oulu, Finland |
| Eeva Kangasniemi | Finnish Clinical Biobank Tampere / University of Tampere / Pirkanmaa Hospital District, Tampere, Finland |

| | |
|---|---|
| Sami Heikkinen | Biobank of Eastern Finland / University of Eastern Finland / Northern Savo Hospital District, Kuopio, Finland |
| Arto Mannermaa | Biobank of Eastern Finland / University of Eastern Finland / Northern Savo Hospital District, Kuopio, Finland |
| Eija Laakkonen | Central Finland Biobank / University of Jyväskylä / Central Finland Health Care District, Jyväskylä, Finland |
| Juha Kononen | Central Finland Biobank / University of Jyväskylä / Central Finland Health Care District, Jyväskylä, Finland |

**Sample Collection Coordination**

| | |
|---|---|
| Anu Loukola | Hospital District of Helsinki and Uusimaa, Helsinki, Finland |

**Sample Logistics**

| | |
|---|---|
| Päivi Laiho | THL Biobank / The National Institute of Health and Welfare Helsinki, Finland |
| Tuuli Sistonen | THL Biobank / The National Institute of Health and Welfare Helsinki, Finland |
| Essi Kaiharju | THL Biobank / The National Institute of Health and Welfare Helsinki, Finland |
| Markku Laukkanen | THL Biobank / The National Institute of Health and Welfare Helsinki, Finland |
| Elina Järvensivu | THL Biobank / The National Institute of Health and Welfare Helsinki, Finland |
| Sini Lähteenmäki | THL Biobank / The National Institute of Health and Welfare Helsinki, Finland |
| Lotta Männikkö | THL Biobank / The National Institute of Health and Welfare Helsinki, Finland |
| Regis Wong | THL Biobank / The National Institute of Health and Welfare Helsinki, Finland |

**Registry Data Operations**

| | |
|---|---|
| Kati Kristiansson | THL Biobank / The National Institute of Health and Welfare Helsinki, Finland |
| Hannele Mattsson | THL Biobank / The National Institute of Health and Welfare Helsinki, Finland |
| Susanna Lemmelä | Institute for Molecular Medicine Finland, HiLIFE, University of Helsinki, Finland |
| Tero Hiekkalinna | THL Biobank / The National Institute of Health and Welfare Helsinki, Finland |
| | Manuel González Jiménez THL Biobank / The National Institute of Health and Welfare Helsinki, Finland |

**Genotyping**

| | |
|---|---|
| Kati Donner | Institute for Molecular Medicine Finland, HiLIFE, University of Helsinki, Finland |

**Sequencing Informatics**

| | |
|---|---|
| Priit Palta | Institute for Molecular Medicine Finland, HiLIFE, University of Helsinki, Finland |
| Kalle Pärn | Institute for Molecular Medicine Finland, HiLIFE, University of Helsinki, Finland |
| Javier Nunez-Fontarnau | Institute for Molecular Medicine Finland, HiLIFE, University of Helsinki, Finland |

**Data Management and IT Infrastructure**

| | |
|---|---|
| Jarmo Harju | Institute for Molecular Medicine Finland, HiLIFE, University of Helsinki, Finland |
| Elina Kilpeläinen | Institute for Molecular Medicine Finland, HiLIFE, University of Helsinki, Finland |
| Timo P. Sipilä | Institute for Molecular Medicine Finland, HiLIFE, University of Helsinki, Finland |
| Georg Brein | Institute for Molecular Medicine Finland, HiLIFE, University of Helsinki, Finland |
| | Oluwaseun Alexander Dada Institute for Molecular Medicine Finland, HiLIFE, University of Helsinki, Finland |
| Ghazal Awaisa | Institute for Molecular Medicine Finland, HiLIFE, University of Helsinki, Finland |
| Anastasia Shcherban | Institute for Molecular Medicine Finland, HiLIFE, University of Helsinki, Finland |
| Tuomas Sipilä | Institute for Molecular Medicine Finland, HiLIFE, University of Helsinki, Finland |

**Clinical Endpoint Development**

| | |
|---|---|
| Hannele Laivuori | Institute for Molecular Medicine Finland, HiLIFE, University of Helsinki, Finland |
| Aki Havulinna | Institute for Molecular Medicine Finland, HiLIFE, University of Helsinki, Finland |
| Susanna Lemmelä | Institute for Molecular Medicine Finland, HiLIFE, University of Helsinki, Finland |
| Tuomo Kiiskinen | Institute for Molecular Medicine Finland, HiLIFE, University of Helsinki, Finland |

**Trajectory Team**

| | |
|---|---|
| Tarja Laitinen | Pirkanmaa Hospital District, Tampere, Finland |
| Harri Siirtola | University of Tampere, Tampere, Finland |

| | |
|---|---|
| Javier Gracia Tabuenca | University of Tampere, Tampere, Finland |

**Biobank Directors**

| | |
|---|---|
| Lila Kallio | Auria Biobank, Turku, Finland |
| Sirpa Soini | THL Biobank, Helsinki, Finland |
| Jukka Partanen | Blood Service Biobank, Helsinki, Finland |
| Kimmo Pitkänen | Helsinki Biobank, Helsinki, Finland |
| Seppo Vainio | Northern Finland Biobank Borealis, Oulu, Finland |
| Kimmo Savinainen | Tampere Biobank, Tampere, Finland |
| Veli-Matti Kosma | Biobank of Eastern Finland, Kuopio, Finland |
| Teijo Kuopio | Central Finland Biobank, Jyväskylä, Finland |
